# Interpretable Machine Learning Reveals Integrated Water Chemistry and Parameter-Specific Nonlinear Responses Shaping *Legionella* spp. and *Mycobacterium* spp. in Drinking Water

**DOI:** 10.64898/2026.04.23.26351579

**Authors:** Jinhao Yang, Huanqi He, Samantha DiLoreto, Kaiqin Bian, Jacob R. Phaneuf, Patrick Milne, Kelsey Pieper, Aron Stubbins, Ching-Hua Huang, Katherine E. Graham, Christopher A. Impellitteri, Ameet Pinto

## Abstract

Traditionally, studies have explored the impacts of individual water chemistry parameters on the persistence of *Mycobacterium* spp. and *Legionella* spp. in isolation with the underlying assumption that these associations are likely monotonic in nature. Yet chemical and microbiological changes are complex, and associations are likely highly combinatorial. In this study, we use interpretable machine learning models to disentangle the integrative and nonlinear associations between water chemistry and occurrence/abundance of *Mycobacterium* spp. and *Legionella* spp. Seasonal data from source water, point-of-entry and distribution systems of eight full-scale drinking water systems demonstrated that shifts in overall water chemistry were associated with the changes in microbial abundance during treatment and distribution. Machine learning models indicated moderate predictive ability of integrated water chemistry towards *Legionella* spp. abundance and towards the occurrence of both *Legionella* spp. and *Mycobacterium* spp., whereas predictive performance for *Mycobacterium* spp. abundance was limited. The association between nitrate and *Legionella* spp. abundance was disinfectant regimes dependent, while dissolved organic carbon exhibited a concentration dependent response type (i.e., positive and negative association). In chloraminated systems, *Legionella* spp. abundance was positively associated with ammonia and nitrate, highlighting the critical role of nitrification. Here, it appears that pH likely influences the initial colonization of *Legionella* spp. while ammonia governs its abundance in drinking water. Overall, this study demonstrates that integrated water chemistry and parameter-specific nonlinear effects collectively explain persistence of *Mycobacterium* spp. and *Legionella* spp. in drinking water systems.

**Synopsis:** This study elucidates the integrative impact of water chemistry and the nonlinear responses of individual water chemistry parameters on the occurrence and abundance of *Mycobacterium* spp. and *Legionella* spp. in drinking water using interpretable machine learning.

**TOC:** 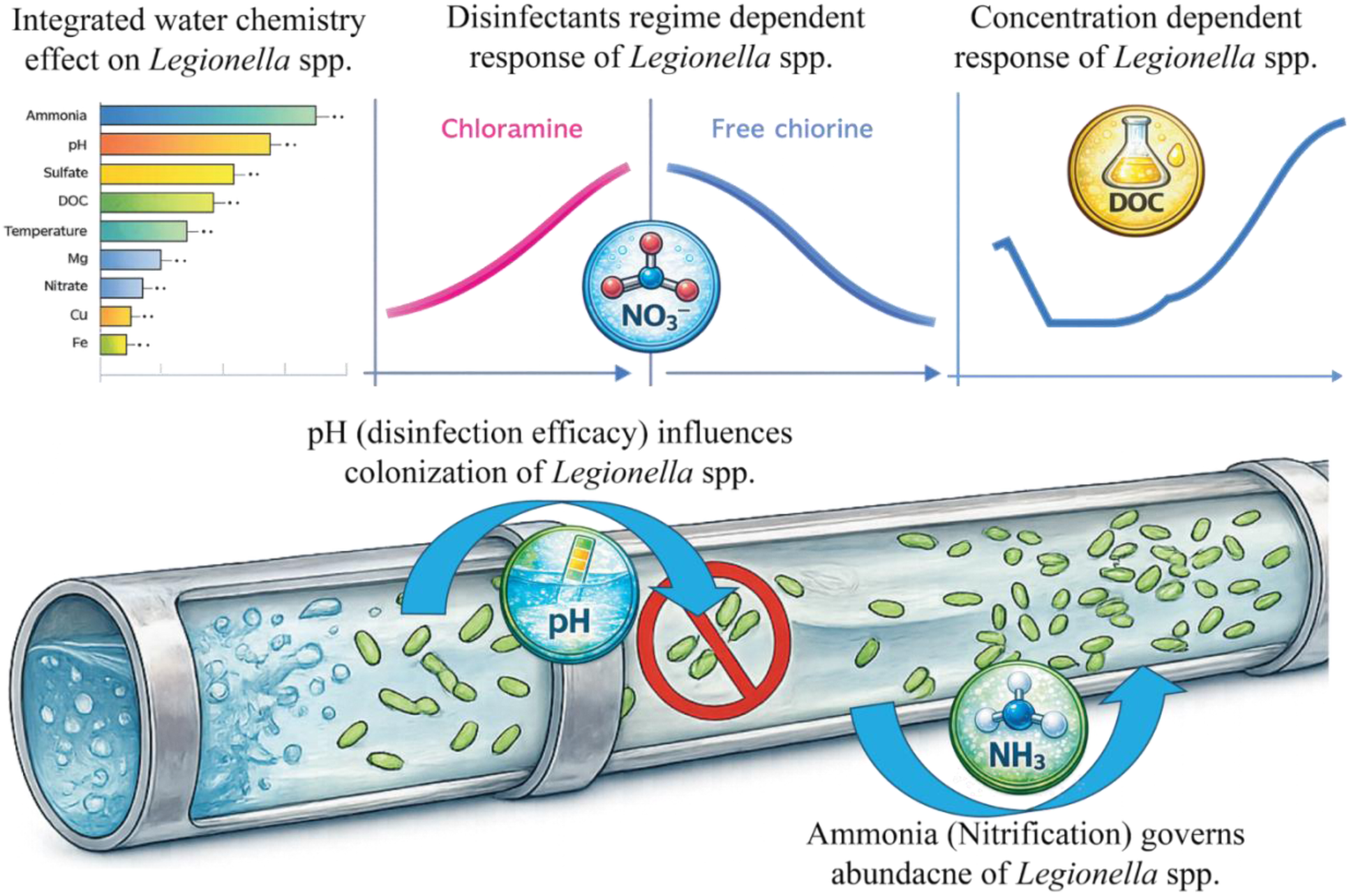

## Introduction

*Mycobacterium* spp. and *Legionella* spp. are widespread in drinking water distribution systems (DWDS). Nontuberculous mycobacteria (NTM) comprises approximately 200 identified species^1,2^, two-thirds of which are capable of infecting humans and animals^2,3^. More than 66 *Legionella* species encompassing over 70 serogroups have been identified^4^, with *Legionella pneumophila* (*L. pneumophila*) accounting for approximately 85–90% of reported infection cases^5^. Infections associated with these bacteria pose significant public health concerns and impose substantial economic burdens, with estimated annual direct healthcare costs of approximately $402 million for Legionnaires’ disease and $11.53 billion for NTM infection in the United States alone^6^. Reported infection incidence for both *L. pneumophila*^7,8^ and NTM^9,10^ has increased in recent years, a trend partly linked to their persistence and proliferation within premise plumbing and DWDS^11–16^. Both *Mycobacterium* spp. and *Legionella* spp. exhibit resistance to commonly applied disinfectants^17,18^, enabling survival, colonization, and regrowth within DWDS. Despite the growing healthcare burden associated with *Mycobacterium* spp. and *Legionella* spp., regulatory strategies specifically targeting these organisms in drinking water remain limited; however, some jurisdictions, such as New Jersey^19^, have adopted more specific building- and system-level risk-management requirements for *Legionella* spp.

Water chemistry is a critical determinant of microbial persistence in drinking water. Previous studies have demonstrated that residual disinfectants^3,20,21^, temperature^22–26^, assimilable organic carbon (AOC)^27–31^ and ammonia^32^ impact the persistence of *Mycobacterium* spp. and *Legionella* spp. However, these associations have frequently been evaluated using correlation analyses^22,23,27,33^ which assume linear or monotonic relationships. Experimental and field-based evidence, however, suggest that microbial responses to physicochemical parameters may instead exhibit nonlinear and combinatorial behavior^22,24,27,28^. Such nonlinear patterns imply that ecological thresholds or range-dependent transitions may be overlooked when relying solely on correlation-based approaches. Beyond assumptions of linearity, correlation-based methods typically evaluate physicochemical parameters individually, implicitly treating each parameter as acting independently. Some studies have also attempted to isolate the influence of specific parameters using pairwise comparisons^3,20^ or simplified laboratory-scale experiments^22^. However, conclusions derived from these approaches may be confounded by concurrent water chemistry interactions^18,34–36^, obscuring the true influence of individual parameters. A recent review synthesizing 117 field- and laboratory-based studies has demonstrated the disinfection efficacy of residual disinfectants against *Legionella* spp. was not determined solely by disinfectant concentration but was influenced by interacting physicochemical parameters such as pH, total organic carbon, and dissolved oxygen^18^. This complexity underscores the challenge of disentangling how integrated water chemistry conditions and parameter-specific response patterns jointly shape their persistence in drinking water.

Machine learning provides a framework to address these limitations by evaluating water chemistry as a multivariate system rather than as a set of independent physicochemical parameters^37–40^. Algorithms such as random forest (RF) ^41–44^, extreme gradient boosting (XGB)^45–47^, and support vector machines (SVM)^48–50^ incorporate multiple parameters simultaneously and assess their relative contributions to microbial persistence. When coupled with interpretable tools such as accumulated local effects (ALE) analyses, these models enable characterization of nonlinear, parameter-specific response patterns while accounting for concurrent chemical conditions^51^. Such approaches allow for direct evaluation of how integrated water chemistry and individual nonlinear effects shape microbial persistence.

In this study, we utilized interpretable machine learning to systematically evaluate the roles of integrated water chemistry and individual physicochemical parameters in shaping *Mycobacterium* spp. and *Legionella* spp. dynamics in full-scale drinking water distribution systems. The objectives of this study were to: (1) assess associations between multivariate water chemistry with both microorganisms’ occurrence and abundance; and (2) characterize nonlinear response patterns of key physicochemical parameters.

## Materials and Methods

### Sample collection and DNA extraction

A total of 182 water samples were collected from eight full-scale drinking water utilities (Utilities 2–9) across diverse geographic regions of the United States^52^. Sampling included source water (n = 35 samples), and finished water at the point of entry (POE, n = 35 samples) and in the drinking water distribution system (DWDS, n = 112 samples) locations representing a range of water ages. Source water included surface water (n = 4 utilities), groundwater (n = 2 utilities), and blended sources (n = 2 utilities), while residual disinfectants were free chlorine (n = 3 utilities) or chloramine (n = 5 utilities) (Table S1). Four sampling campaigns were conducted at each utility between January and December 2024. For microbial analyses, 500 mL of source water and 2,000 mL of finished water (POE and DWDS) were collected in sterile Nalgene™ PPCO bottles (Thermo Fisher Scientific, MA, USA) pre-dosed with 10 mg sodium thiosulfate to neutralize residual disinfectants. Additionally, 1,000 mL water was collected at each site in acid-washed HDPE bottles for dissolved organic matter analysis, and 125 mL was collected in HDPE bottles for anion analysis. All samples were shipped on ice via overnight delivery to the laboratory. Upon arrival, samples were filtered through enclosed Sterivex™ 0.22 μm filter units (Millipore) and stored at −20 °C prior to DNA extraction. DNA was extracted using the DNeasy PowerWater Kit (Qiagen) following the manufacturer’s protocol with minor modifications as previously described^53^. DNA yield was quantified on the Qubit Flex fluorometer (Invitrogen, CA, USA) using the 1× Qubit dsDNA High-Sensitivity Assay Kit (Invitrogen, CA, USA) following the manufacturer’s instructions (Table S2).

### Water Quality and Microbial Analyses

Free chlorine, total chlorine, water temperature, and pH were measured and reported by each water utility. Ammonia was quantified using Hach Method 10023 (Hach Company). Dissolved organic carbon (DOC) was analyzed using the Shimadzu TOC-L+TN analyzer. Anions, including nitrite, nitrate, chloride, sulfate, and fluoride were measured by ion chromatography (930 Compact IC Flex, Metrohm, Herisau, Switzerland) following EPA Method 300.1 (Parts A and B). Metal elements (Cu, Zn, Mg, Na, Al, K, Ca, and Fe) and phosphorus (P) were quantified by inductively coupled plasma mass spectrometry (ICP-MS). Total and intact cell counts were determined by flow cytometry as previously described^54^. *L. pneumophila* and *Pseudomonas aeruginosa* (*P. aeruginosa*) were quantified using IDEXX Legiolert and Pseudalert assays (100 mL format), respectively, and results were reported as MPN/100 mL. Heterotrophic plate counts (HPC) were determined using IDEXX SimPlate for HPC (100 mL format).

Gene copies of *Legionella* spp. (23S rRNA gene)^55^, *L. pneumophila* (mip gene)^55^ were quantified using duplex digital PCR (dPCR), while *Mycobacterium* spp. (16S rRNA gene)^56,57^ and *Mycobacterium avium* (*M. avium*, 16S rRNA gene)^57,58^ were quantified using singleplex dPCR. Assays were performed according to published protocols^55,56,57,58^, with thermocycling conditions and primer and probe concentrations optimized to improve specificity and performance. Primer and probe sequences, gBlock standards, and thermocycling conditions are provided in the Supporting Information (Table S3). The limit of blank (LOB), limit of detection (LOD), and limit of quantification (LOQ) were determined using a previously published method^59^ and are summarized in Table S4. All reactions were performed on a QIAcuity™ Digital PCR System (QIAGEN, Hilden, Germany) using 8.5k Nanoplates (cat.no. 250021) according to the manufacturer’s instructions. Reagent composition and volumes for the 12 μL dPCR reactions used in each assay are listed in Table S5. DNA extracts, no-template controls (sterile Nanopure water), and positive controls (synthetic gBlock gene fragments) were included in triplicate for each run. Thresholds were set manually for each assay based on fluorescence separation between positive and negative partition clusters, using no-template controls and positive controls as references. Samples exhibiting inhibition were diluted 10-fold in DNase-free water and re-quantified. Samples with concentrations greater than LOQ/2 were classified as positive and retained at their measured values, whereas samples with concentrations at or below LOQ/2 were classified as negative and assigned a value of zero.

### Machine Learning Modeling

The associations of integrated water chemistry with the occurrence and log10-transformed abundance of *Legionella* spp. and *Mycobacterium* spp. in finished drinking water (i.e., POE and DWDS) were evaluated using a suite of machine learning approaches in R^60^. Species-level models for *M. avium* and *L. pneumophila* were not developed due to a high proportion of non-detects (>70%) resulting in insufficient non-zero data and limited data variation for reliable modeling. Candidate predictors included measured water chemistry parameters previously reported to be associated with these microorganisms and exhibiting limited missing values. These parameters include pH, temperature, ammonia, DOC, nitrate, sulfate, Mg, Fe, Cu and Zn. Collinearity among parameters was evaluated using variance inflation factors (VIF), and no high multicollinearity was identified. Across the selected predictors, the number of missing observations per parameter ranged from 1 to 28 out of 147 samples. Missing values were imputed using k-nearest-neighbor (kNN) imputation applied to predictors that were mean-centered and scaled to unit variance prior to imputation. RF, SVM and XGB regression models were implemented using repeated nested cross-validation (outer 5-fold with five repeats; inner 5-fold for hyperparameter tuning) to predict abundance. Performance on held-out outer test folds was evaluated using mean squared error (MSE), root mean squared error (RMSE), and fold-level R^2^. Feature importance was estimated to identify key predictors. Model interpretability was further assessed using ALE computed with the *iml* framework on the final XGB regression model to quantify marginal predictor effects. For occurrence prediction, SVM, RF, and XGB classifiers were evaluated using repeated nested cross-validation (outer 5-fold with five repeats; inner 5-fold for hyperparameter tuning). Predicted probabilities from outer test folds were converted to binary outcomes using thresholds determined by the Youden index. Model performance was summarized by Area Under the Curve (AUC), sensitivity, specificity, balanced accuracy, and F1 score. Receiver Operating Characteristic (ROC) significance was assessed against a null classifier with constant probability (0.5).

### Statistical analysis

Parametric one-way analysis of variance or nonparametric Kruskal–Wallis tests were conducted to compare physicochemical parameters and microbial abundances among sample types and disinfectant regimes, depending on normality and variance assumptions. Post-hoc pairwise comparisons were conducted with Bonferroni-adjusted P values. Statistical significance was defined at P < 0.05. Principal component analysis (PCA) was performed to assess multivariate differences in water chemistry. Permutational multivariate analysis of variance (PERMANOVA) was conducted to test for significant compositional differences based on Euclidean distance matrices. Multivariate dispersion among groups was evaluated using permutational analysis of multivariate dispersion (PERMDISP). Statistical significance was assessed at 999 permutations. Spearman’s rank correlation analysis was used to evaluate monotonic associations between *Legionella* spp. abundance and water chemistry parameters. All statistical analyses were conducted in R^60^.

## Results and Discussion

### Changes in water chemistry were associated with changes in microbial abundance during water treatment and distribution

PCA was used to assess differences in overall water chemistry among source water, POE, and DWDS water. The first two principal components explained 42.9% of the total variance between source water and POE (**Figure 1A**). Clear compositional separation was observed and confirmed by PERMANOVA (R² = 0.10, P < 0.05), indicating substantial chemical alteration during treatment. Several parameters, including Ca, Al, Cu, Fe, Zn, P, pH, and fluoride differed significantly between source water and POE (**Figure S1 and S2**, P < 0.05). However, treated water chemistry remained relatively stable during distribution, as evidenced by tight clustering of POE and DWDS samples in the PCA (**Figure 1B**) and the absence of significant differences between them based on PERMANOVA analyses (P > 0.05). Consistent with this pattern, most individual parameters, except for Zn, Cu, and Fe, did not differ significantly between POE and DWDS samples (**Figure S1-2,** P > 0.05), indicating that the treated water chemistry was largely preserved across the distribution system. Although multivariate dispersion differed among sample types (PERMDISP, P < 0.05), pairwise comparisons showed that POE and DWDS exhibited lower dispersion than source water (P < 0.05; **Figure 1C**), while no significant difference was observed between POE and DWDS (P > 0.05). These results indicate that water treatment reduced the variability in chemical composition across utilities, and this remained consistent through the distribution systems.

**Figure 1.**
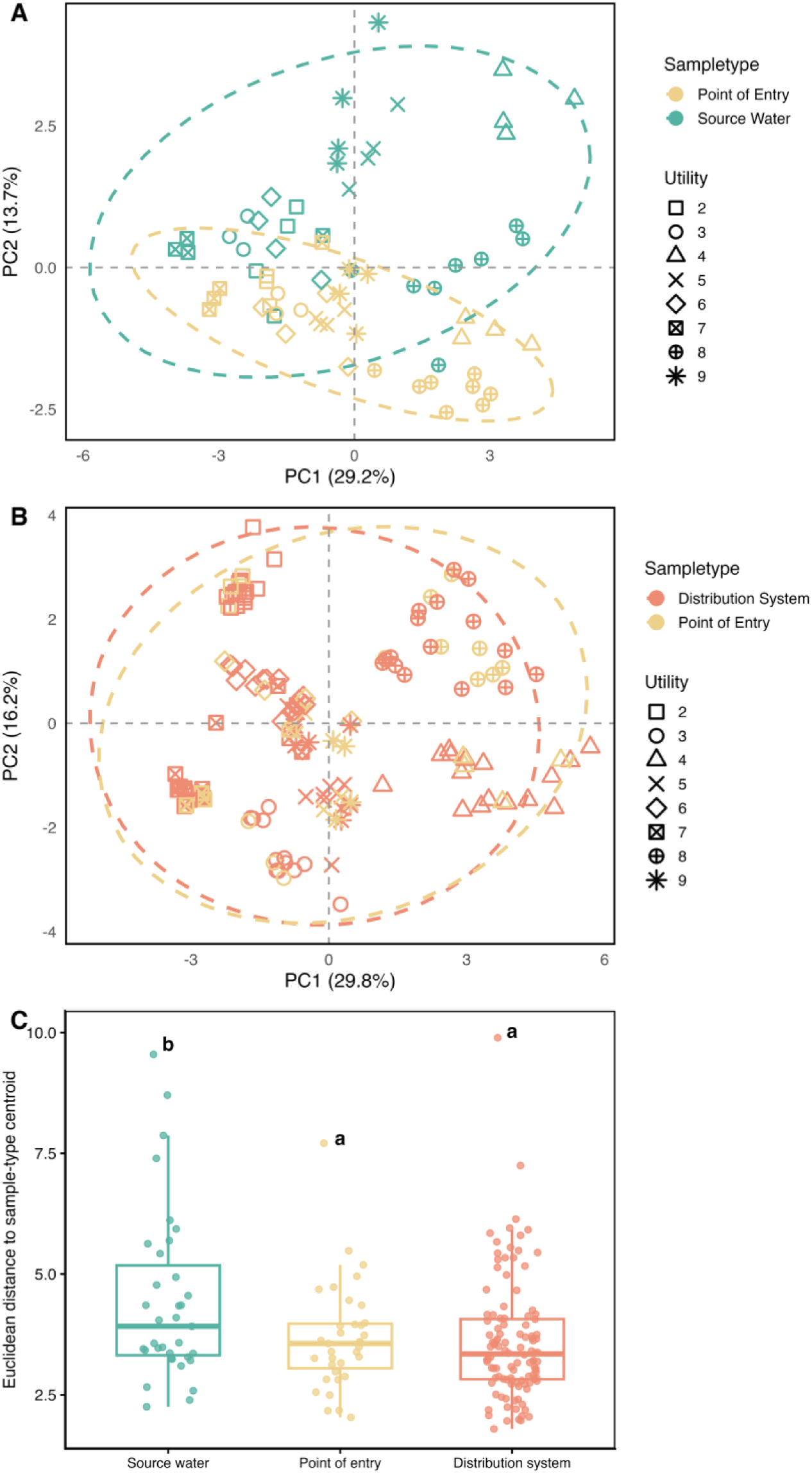
Principal component analysis of water chemistry comparing (A) source water and point-of-entry (POE) samples and (B) POE and drinking water distribution system (DWDS) samples. (C) Multivariate dispersion of water chemistry based on Euclidean distances to sample-type centroids assessed using permutational analysis of multivariate dispersions; different letters (a and b) indicate significant differences among sample types (P < 0.05).

Consistent with these chemical shifts, bulk microbial indicators demonstrated pronounced reduction post-treatment. Total cell counts in source water averaged 5.36 ± 0.67 log10 cells mL⁻¹, which were 1.22-fold and 1.19-fold higher than POE and DWDS, respectively (**Figure 2A**, P < 0.05). Intact cell counts in source water were 4.99 ± 0.97 log10 cells/mL, corresponding to 1.70-fold and 1.67-fold higher levels than POE and DWDS, respectively (**Figure 2B**, P < 0.05). The HPC in source water (2.28 ± 1.11 log10 CFU/mL) was 3.86 and 3.08 times higher than POE and DWDS, respectively (**Figure 2C**, P < 0.05). However, none of these microbial indicators differed significantly between POE and DWDS (P > 0.05), indicating microbial concentrations remained largely stable during distribution. This stability was likely maintained by the persistent residual disinfectants, with measured free chlorine and total chlorine concentrations of 1.13 ± 0.57 and 2.56 ± 0.76 mg/L in chlorinated and chloraminated DWDS samples, respectively. Similar trends were observed for *P. aeruginosa* abundance quantified by Pseudalert assays, which averaged 1.74 ± 1.60 log10 MPN/100 mL in source water and was 98.3 times and 310.7 times higher than that in POE and DWDS, respectively (**Figure 2D**, P < 0.05). In contrast, the Pseudalert-based *P. aeruginosa* concentrations did not differ significantly between POE and DWDS (P > 0.05).

**Figure 2.**
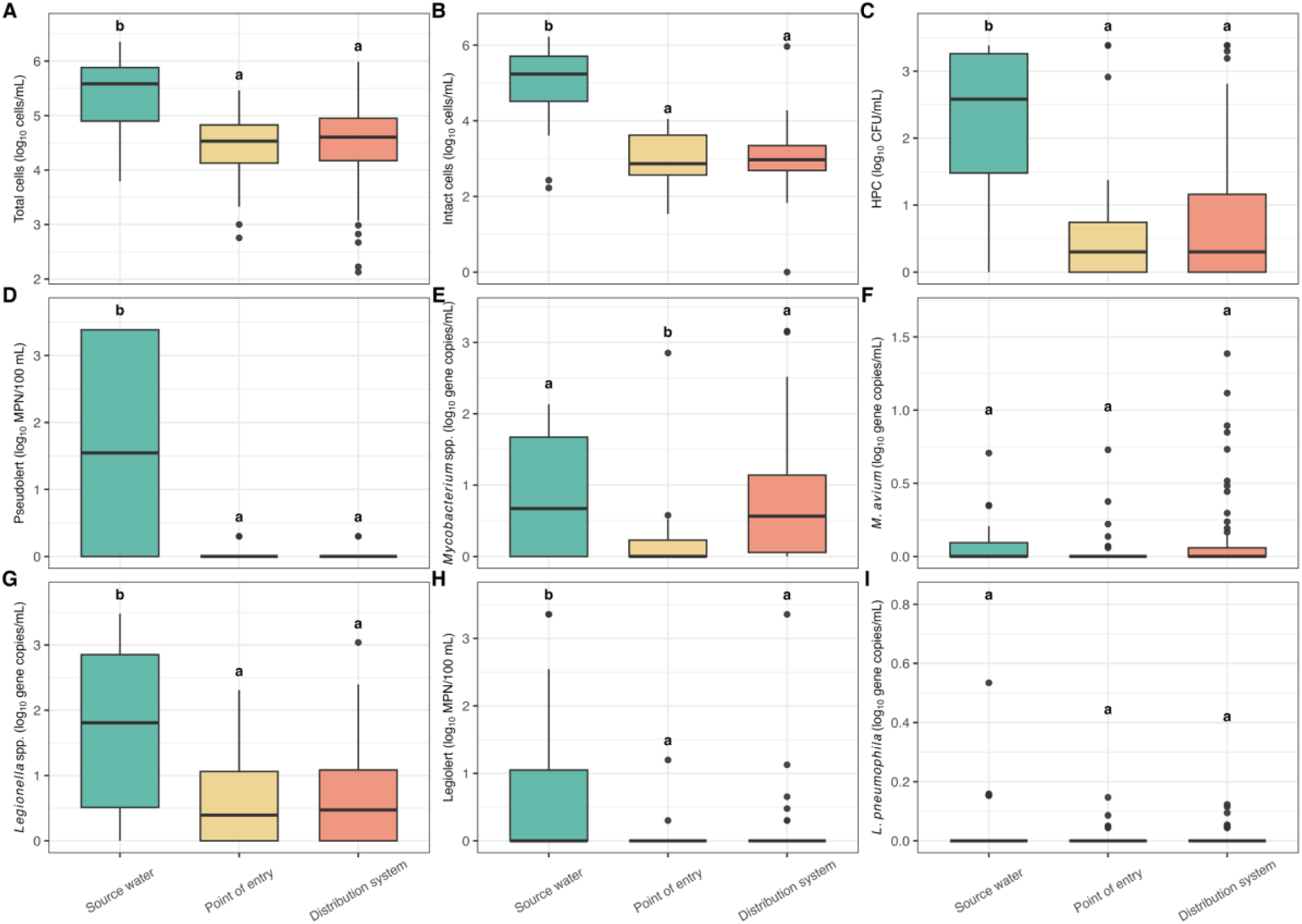
Microbial dynamic across source water, point-of-entry, and drinking water distribution system samples. (A) Total cells, (B) intact cells, (C) heterotrophic plate counts, (D) Pseudalert-based *Pseudomonas aeruginosa*, (E) *Mycobacterium* spp., (F) *M. avium*, (G) *Legionella* spp., (H) Legiolert-based *L. pneumophila*, and (I) *L. pneumophila*. Different letters (a and b) above boxplots denote statistically significant differences among sample types (P < 0.05).

The frequency of detection (FOD) of *Mycobacterium* spp. in source water across the eight utilities was 61.0% ± 39.7% and decreased to 47.9% ± 35.8% at POE. In DWDS samples, the average FOD increased to 80.0% ± 15.1%, which was 1.67-fold higher than that observed at POE. The gene copies of *Mycobacterium* spp. in source water were 0.78 ± 0.79 log10 gene copies/mL, which were 3.20 and 1.08 times higher than POE and DWDS samples, respectively (**Figure 2E**, P < 0.05). In addition, gene copies of *Mycobacterium* spp. in DWDS (0.72 ± 0.72 log10 gene copies/mL) were 3.0 times higher than at POE (P < 0.05). Together, these patterns indicate substantial reduction of *Mycobacterium* spp. during water treatment followed by significant increase during distribution, which may be attributed to regrowth and/or biofilms detachment. The FOD of *M. avium* in source water and POE across the eight utilities was 32.3% ± 31.9% and 20.3% ± 21.1%, respectively. In DWDS samples, the average FOD increased slightly to 25.4% ± 18.3%. Gene copies of *M. avium* ranged from below LOD to 1.38 log10 gene copies mL⁻¹ and did not differ significantly between source water, POE, and DWDS (**Figure 2F**, P > 0.05), indicating relatively low and stable abundance throughout water treatment and distribution.

The FOD of *Legionella* spp. in source water across the eight utilities was 90.6% ± 18.6% and decreased to 70.3% ± 32.7% at POE, with a slight increase to 74.8% ± 27.1% in DWDS. The gene copies of *Legionella* spp. in source water were 1.66 ± 1.19 log10 gene copies/mL, which were 2.55 and 2.39 times higher than POE and DWDS, respectively (**Figure 2G**, P < 0.05). However, no significant difference was observed between POE and DWDS (P > 0.05), indicating effective reduction during treatment but relative stability during distribution. Notably, the abundance dynamic patterns during treatment and distribution of *Legionella* spp. differed from those observed for *Mycobacterium* spp. where the *Mycobacterium* spp. abundance decreased during treatment and then increased during distribution, suggesting distinct ecological patterns and potential differences in regrowth dynamics. Both culture-based Legiolert and molecular dPCR targeting the *mip* gene were used to quantify *L. pneumophila* (**Figure 2H and I**). Across the eight utilities, the FOD of *L. pneumophila mip* gene and *L. pneumophila* detected by Legiolert in source water averaged 19.8% ± 22.7% and 37.7% ± 32.6%, respectively, and declined to 16.1% ± 26.8% and 6.25% ± 10.8% at POE. In DWDS samples, the corresponding FODs further decreased slightly to 14.5% ± 16.2% and 5.71% ± 5.34%, respectively. Legiolert-based *L. pneumophila* abundances in source water averaged 0.67 ± 1.02 log10 MPN /100 mL and was 16.8-fold and 11.2-fold higher than POE and DWDS, respectively (P < 0.05). In contrast, *L. pneumophila* gene copies did not differ significantly between source water and DWDS samples (P > 0.05). Collectively, these findings indicate that culturable *L. pneumophila* was substantially reduced post water treatment, whereas *L. pneumophila* gene copies remained comparatively stable which is likely due to methodological differences between DNA-based detection and culture-based assays. Additionally, neither *L. pneumophila mip* gene abundance nor Legiolert-based *L. pneumophila* abundance differed significantly between POE and DWDS samples (P > 0.05), indicating relatively stable *L. pneumophila* levels during the distribution. The overall correspondence between shifts in multivariate water chemistry profiles and microbial patterns suggests that integrated water chemistry was closely associated with microbial dynamics during treatment and distribution.

### *Mycobacterium* spp. and *Legionella* spp. concentrations in finished drinking water demonstrated contrasting associations with source water concentrations

In finished drinking water, samples supplied by groundwater exhibited higher gene copies of both *Mycobacterium* spp. (0.69 ± 0.69 log10 gene copies mL⁻¹) and *Legionella* spp. (0.91 ± 0.71 log10 gene copies mL⁻¹) than those supplied by surface water, corresponding to 1.57-fold and 2.12-fold higher concentration, respectively (**Figure 3A**, P < 0.05). In contrast, *Mycobacterium* spp. abundance in surface water (1.14 ± 0.82 log10 gene copies mL⁻¹) was 4.62-fold higher than in groundwater (**Figure 3B**, P < 0.05). Similarly, *Legionella* spp. averaged 1.93 ± 1.13 log10 gene copies mL⁻¹ in surface water, 1.85-fold higher than in groundwater (P < 0.05). Together, these results demonstrate that although *Mycobacterium* spp. and *Legionella* spp. abundances were lower in untreated groundwater than in surface water, finished drinking water supplied by groundwater exhibited higher gene copies following treatment and distribution. This inverse pattern suggests that source water type does not solely determine the abundance of *Mycobacterium* spp. and *Legionella* spp. in finished drinking water. Instead, treatment-related processes (e.g., reseeding within treatment infrastructure) and post-treatment processes (e.g., potential regrowth^61–64^ or biofilm detachment^65–68^ during distribution) also play an important role. Utility-specific variations of *Legionella* spp. and *Mycobacterium* spp. abundance were further evaluated for potential evidence of reseeding during treatment. Removal efficacy ranged from 41.9–97.6% for *Legionella* spp. and 23.2–100% for *Mycobacterium* spp., indicating substantial inter-utility variability in treatment performance. Notably, gene copies of these two microorganisms in POE from several groundwater-suppled utilities exceeded those detected in the corresponding source water, including *Legionella* spp. at utility 9, and *Mycobacterium* spp. at utilities 8 (ground water) and 9 (**Figure 3C**). These patterns may be consistent with reseeding within treatment. However, because these concentrations were below the LOQ, these increases are not sufficient on to confirm reseeding, as analytical uncertainty associated with low-level dPCR quantification may also have contributed to the observed pattern.

**Figure 3.**
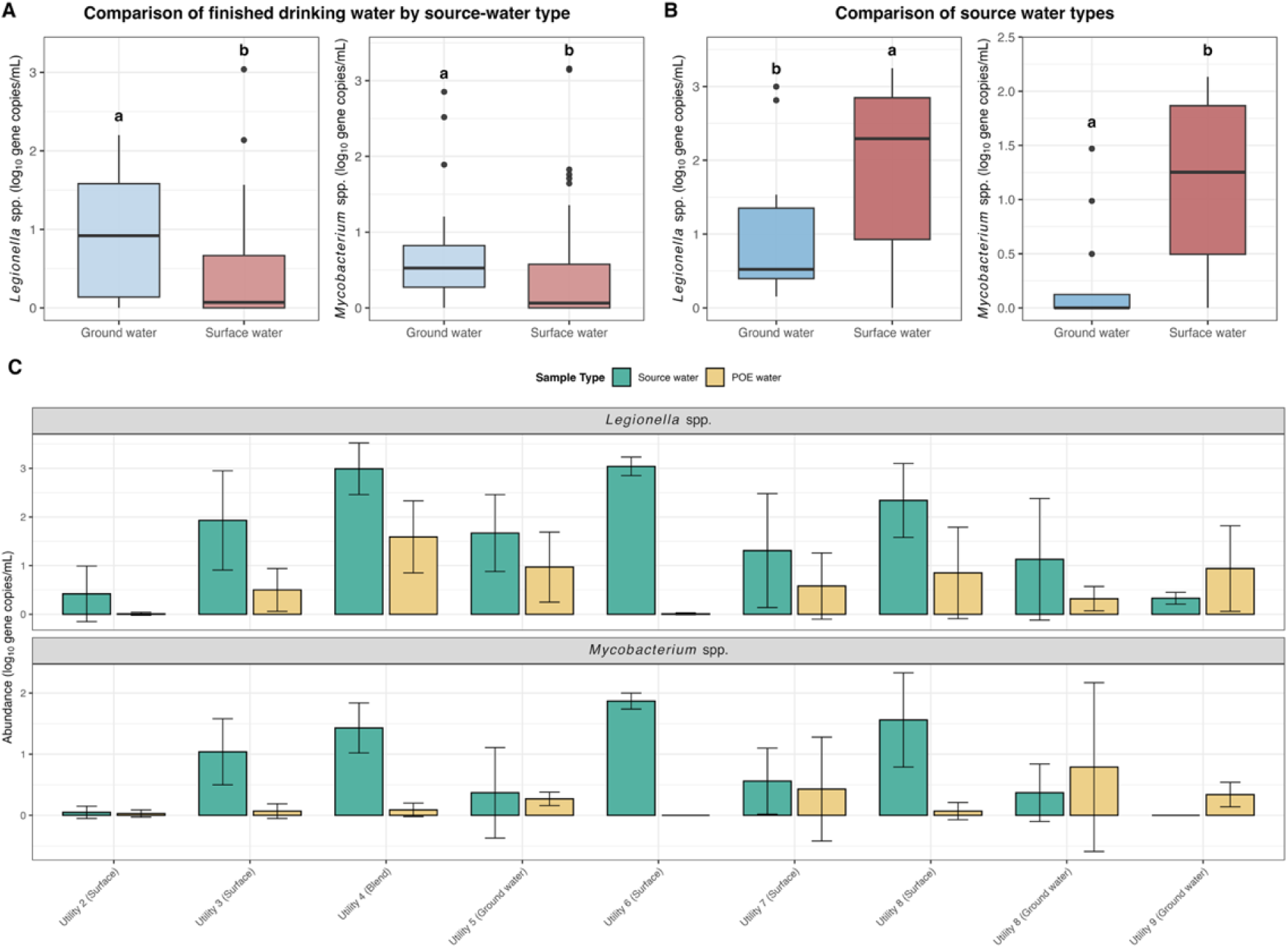
Gene copy of *Legionella* spp. and *Mycobacterium* spp. in (A) finished drinking water supplied by groundwater and surface water sources and (B) source waters by source type. Different letters (a and b) indicate statistically significant differences between source types (P < 0.05). (C) Utility-specific gene copies in source water and corresponding point-of-entry samples.

### Variation in *Mycobacterium* spp. and *Legionella* spp. abundance aligned with disinfectant regime-level differences in water chemistry

The first two PCA axes explained 46.0% of the variance of water chemistry in finished water, and chloraminated and chlorinated samples formed distinct clusters (PERMANOVA: R² = 0.122, P < 0.05) (**Figure 4A**). Higher concentrations/levels of ammonia, pH, and DOC were measured in chloraminated waters relative to chlorinated systems (**Figure S3,** P < 0.05). In contrast, nitrate and sulfate concentrations were significantly higher in chlorinated waters relative to chloraminated systems (**Figure S3,** P < 0.05). Several additional physicochemical parameters, including chloride, Zn, Al, Ca, Cu, Fe, K, Mg, P, and Na, differed significantly between disinfectant regimes (P < 0.05**; Figure S3 and S4**). Notably, elevated DOC, nitrate, and sulfate concentrations within either disinfectant regime were consistent with their corresponding levels in source water, and none of these constituents changed significantly during treatment or distribution (**Figure S5**, P > 0.05). This indicates that source water composition, rather than residual disinfectants, primarily drove these differences in finished water. Collectively, these findings demonstrate that chlorinated and chloraminated systems monitored in this study displayed distinct overall chemical regimes shaped by both disinfectant strategy and source water composition.

**Figure 4.**
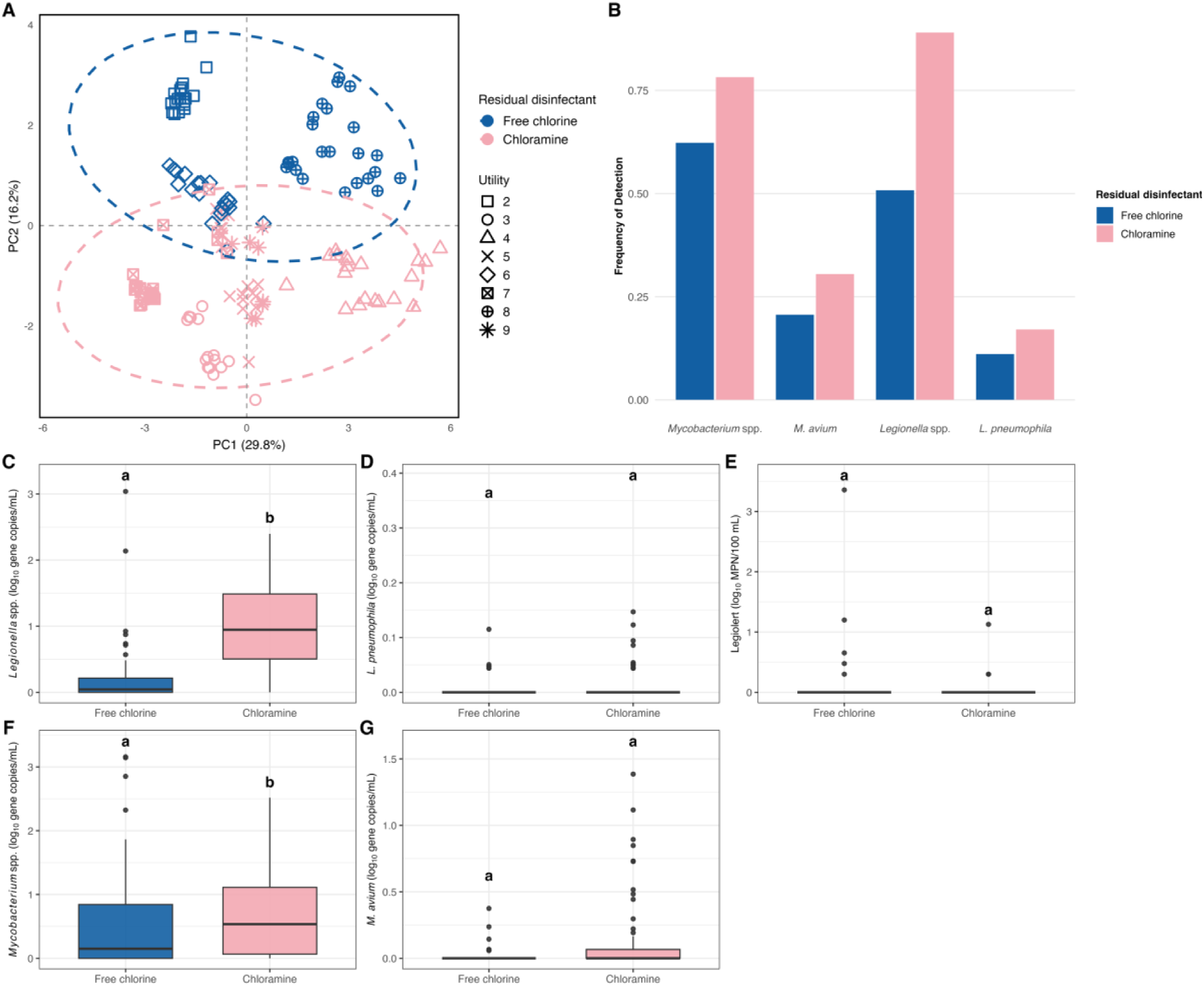
(A) Principal component analysis of water chemistry in point-of-entry and drinking water distribution system samples disinfected with free chlorine or chloramine. (B) Frequency of detection of *Mycobacterium* spp., *M. avium*, *Legionella* spp., and *L. pneumophila* in finished drinking water under free chlorine and chloramine residuals. (C–G) Abundances of (C) *Legionella* spp., (D) *L. pneumophila*, (E) Legiolert, (F) *Mycobacterium* spp., and (G) *M. avium* in chlorinated and chloraminated finished water. Different letters (a and b) indicate statistically significant differences between disinfectant types (P < 0.05).

The FOD of *Mycobacterium* spp., *M. avium*, *Legionella* spp., and *L. pneumophila* in chloraminated finished water (78.2%, 30.5%, 89.0%, and 17.1%, respectively) was greater than those in chlorinated finished water (**Figure 4B**). The gene copies of *Mycobacterium* spp. and *Legionella* spp. in chloraminated finished water were 0.68 ± 0.64 and 1.04 ± 0.71 log10 gene copies/mL, respectively, which were 1.31-fold and 4.73-fold higher than chlorinated finished water (**Figure 4C** and F, P < 0.05). In contrast, the abundances of *M. avium* and *L. pneumophila* were low (0.10 ± 0.26 and 0.01 ± 0.03 log10 gene copies mL⁻¹, respectively) and did not differ significantly between chloraminated and chlorinated finished water (**Figure 4D and G**, P > 0.05). Likewise, Legiolert-based *L. pneumophila* abundance showed no significant differences in finished drinking water between residual disinfectant regimes (**Figure 4E**, P > 0.05). Collectively, chloraminated finished water exhibited higher FOD and greater abundances of *Mycobacterium* spp. and *Legionella* spp. compared with chlorinated finished water, consistent with previous studies^3,69^. This pattern could be ascribed to the lower biocidal strength of chloramine relative to free chlorine^70^ which was evidenced by the significantly higher total cell counts and HPC in chloraminated finished drinking water (**Figure S6**, P < 0.05). Further, higher ammonia concentrations in chloraminated systems may promote nitrification, potentially favoring the persistence of *Mycobacterium* spp. and *Legionella* spp^71,72^.

The alignment between disinfectant regime-level water chemistry differences across multiple parameters and variation in *Mycobacterium* spp. and *Legionella* spp. abundances suggests that disinfectant-specific chemical environments shape microbial dynamics and that no single water chemistry parameter is a critical determinant. The influence of individual physicochemical parameter on microbial dynamics has been widely documented^22–31^. For example, assimilable organic carbon (AOC) has been consistently associated with increased microbial abundance, with positive correlations reported between AOC and *Mycobacterium* spp. abundance in drinking water^27,28^. However, prior studies have largely relied on correlation-based approaches that evaluate parameters independently, comparing correlation coefficients and statistical significance^23,27,28^. Experimental studies have explored interactive effects under controlled conditions. For instance, simulated water heaters equipped with copper or PEX pipe sections were subjected to varying AOC concentrations (0–700 μg/L) and incrementally increased temperature to investigate their combined influence on *L. pneumophila*^22^. Similar experimental designs have been used to examine the interactive effects of corrosion, copper, and chloramines on *Legionella* spp. and *Mycobacterium* spp., systematically introducing cupric ions and chloramine to assess microbial responses^20^. While such studies provide valuable mechanistic insight, they typically focus on a limited number of variables and do not capture the multidimensional, co-varying complexity characteristic of full-scale DWDS. Such approaches did not explicitly account for the integrated nature of water chemistry, in which physicochemical parameters co-vary. Assuming parameter independence may oversimplify system-level behavior, particularly given well-documented interactions among physicochemical parameters^20,22^. For instance, pH, temperature, organic carbon, and nitrogen species strongly influence residual disinfectants efficacy, thereby altering microbial responses under different chemical contexts^34–36,73^. These interdependencies indicate that microbial persistence is impacted not only by the magnitude of individual parameters but also the overall chemical environment. The present findings underscore the importance of evaluating multivariate chemical environments when interpreting microbial abundance dynamics in full-scale DWDS.

### Nonlinear and stage-specific chemical regulation shapes *Legionella* spp. and *Mycobacterium* spp. dynamics in finished drinking water

To evaluate the integrated water chemistry effects on *Legionella* spp. abundance, RF, SVM, and XGB regression models were applied. RF (R² = 0.51 ± 0.30) and XGB (R² = 0.60 ± 0.16) achieved higher predictive performance than SVM, with XGB exhibiting lower variability across cross-validation folds, indicating greater predictive stability. The RMSE of the XGB model for *Legionella* spp. was 0.45 ± 0.12 log10 gene copies/mL, which was small relative to the observed abundance range (0–3.0 log10 gene copies/mL), indicating reasonable predictive accuracy. These metrics indicate that integrated water chemistry collectively provide robust predictive capacity for *Legionella* spp. abundance. Nevertheless, a substantial fraction of variance remained unexplained, likely reflecting the influence of unmeasured factors such as microbial interactions^74–81^, biofilm ecology^66,82^, or additional environmental factors^83–86^ not captured in the model. Permutation importance analysis from the XGB regression model identified ammonia (30.6% ± 9.3%) as the most important predictor of *Legionella* spp. abundance (**Figure 5A**). Nitrate contributed 4.7% ± 1.3%, and collectively nitrogen species accounted for 35.3% of total permutation importance. The ALE plots further reveal nonlinear, parameter-specific responses of ammonia and nitrate within this multivariate context. Predicted *Legionella* spp. abundance increased sharply at high concentration ranges of ammonia (**Figure 5B**), while nitrate displayed a complex, piecewise pattern characterized by two localized ranges of positive marginal effects, each followed by declines at higher concentrations (**Figure 5C**). Notably, model-predicted increases in *Legionella* spp. abundance associated with ammonia and nitrate were predominantly observed in chloraminated finished drinking water. The concurrent positive association of ammonia and nitrate with predicted *Legionella* spp. abundance suggest that ammonia-driven nitrification may be associated with the observed responses. Spearman correlation analysis further demonstrates a strong positive correlation between ammonia and nitrate within chloraminated samples (**Figure S7A**; r = 0.70, P < 0.05). Additional support for the occurrence of nitrification was provided by the detection of nitrite in a subset of chloraminated finished water samples, whereas nitrite was absent in corresponding source waters. Collectively, these observations indicate that nitrification occurred within chloraminated distribution systems and was associated with elevated *Legionella* spp. abundance.

**Figure 5.**
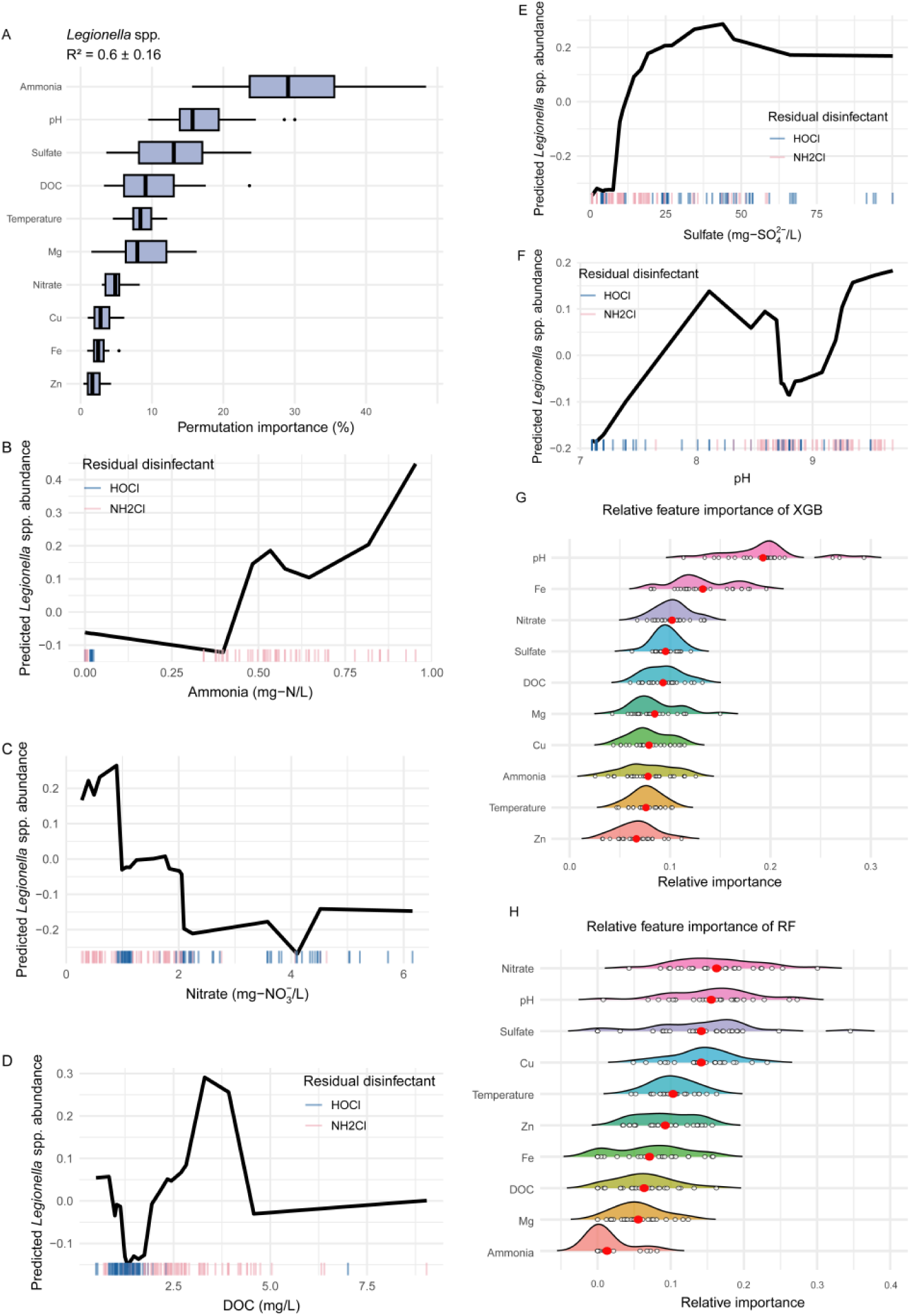
(A) Permutation importance from repeated cross-validation of XGB regression model predicting *Legionella* spp. abundance. (B–F) Accumulated local effects (ALE) curves from the XGB regression model illustrating the marginal effects of the five influential predictors on predicted *Legionella* spp. abundance. The x-axis indicates the observed value of each parameter, and the y-axis indicates the centered ALE effect on predicted *Legionella* spp. abundance; values below zero indicate predictions lower than the mean model prediction. Relative feature importance from the (G)XGB classification model for *Legionella* spp. occurrence and (H) the RF classification model for *Mycobacterium* spp. occurrence; red points indicate mean relative importance across cross-validation runs.

Nitrification generates nitrite as an intermediate, which can react with chloramine and accelerate disinfectant decay^87,88^. In addition, ammonia oxidation may alter monochloramine equilibria, leading to subsequent hydrolysis of monochloramine^89^. Together, these processes would reduce chloramine residual concentrations and diminish the biocidal pressure, facilitating the persistence of *Mycobacterium* spp. and *Legionella* spp. However, the observed positive correlation between ammonia and total chlorine suggests that widespread bulk chloramine decay was unlikely across the sampled systems (**Figure S7B**, r = 0.94, P < 0.05). Therefore, it is reasonable to infer that nitrification likely occurred at localized or biofilm-associated scales rather than uniformly in the bulk water, and large-scale chloramine decay was not the primary mechanism associated with *Legionella* spp. abundance. Beyond chloramine decay, nitrifying bacteria can contribute to both the formation and maintenance of biofilms under carbon-limited conditions typical of DWDS. Previous laboratory-scale studies have revealed that *Nitrosomonas europaea* can support heterotrophic growth and biofilm development through organic carbon production during ammonia oxidation^71^. In addition, sustained ammonia oxidation appears critical for maintaining established biofilms, as inhibition of ammonia oxidation has been shown to cause biofilm destabilization and dispersal^71^. Biofilms provide protected niches that facilitate the survival and regrowth of *Legionella* spp. under chloraminated conditions^72^, and biofilm detachment may further increase their abundance in bulk water^65,66^. Together, these processes provide a plausible ecological mechanism linking ammonia-mediated nitrification to enhanced *Legionella* spp. persistence within the chloraminated chemical regime. In contrast, the decreasing trend observed in the ALE response for nitrate in chlorinated samples suggests a different parameter-specific effect under an alternative chemical regime. This negative association may be related to intensified competition for limited AOC under nitrate-enriched conditions. Previous studies have demonstrated the widespread presence of facultative denitrifying bacteria in drinking water biofilms and their capacity to reduce nitrate under favorable redox conditions^90,91^. Although denitrification activity was not directly measured here, nitrate-supported heterotrophic metabolism could increase competition for limited AOC^90^. Such competition may constrain *Legionella* spp. proliferation, providing a plausible mechanistic explanation for the observed negative nitrate–response pattern observed in chlorinated distribution systems.

The high importance of DOC (9.9% ± 4.9%) aligned with prior work demonstrating that organic carbon availability was a key determinant of biological stability in finished drinking water. DOC displayed a biphasic response pattern across concentration ranges. The predicted *Legionella* spp. response declined slightly at low DOC concentration (**Figure 5D**), consistent with carbon-limited conditions that may favor fast-growing heterotrophs over *Legionella* spp. As DOC increased, the predicted response rose sharply, suggesting the increased organic carbon availability could promote *Legionella* spp. proliferation once carbon limitation was alleviated. Sulfate accounted for 12.7% ± 5.6% and exhibited a monotonic positive association in ALE analysis (**Figure 5E**). ALE analysis indicates that increasing sulfate concentrations were associated with elevated predicted *Legionella* spp. abundance within both disinfectant regimes. However, sulfate concentrations were significantly higher in chlorinated systems, where overall *Legionella* spp. abundance was generally lower than in chloraminated systems. This contrast suggests that sulfate was not a primary system-level driver but rather exerted minor positive influence within each disinfectant regime. The positive association between sulfate and *Legionella* spp. abundance may reflect redox-mediated processes occurring within biofilms. Sulfate can function as an alternative electron acceptor for sulfate-reducing bacteria under anaerobic niches provided by biofilms and corrosion tubercles^92,93^. Activation of sulfate reduction would exacerbate localized corrosion^92–95^, which would enhance *Legionella* spp. regrowth by providing extensive surface area for biofilm attachment^96^ and releasing nutrients^97^. The contribution of pH towards the predictive performance was 17.3% ± 5.2%, with progressively higher pH associated with increased predicted *Legionella* spp. abundance (**Figure 5F**).

To determine whether water chemistry exerted distinct roles in affecting *Legionella* spp. occurrence (presence/absence) as compared to abundance, RF, SVM, and XGB classification models were applied to evaluate predictors of occurrence. ROC curves aggregated across all nested cross-validation runs indicated good classification performance for all three models, as reflected by their consistently elevated true-positive rates across the range of false-positive rates (**Figure S8**). Among the models, RF showed the highest significant-run ratio (22/25, 88%), followed closely by XGB (21/25, 84%), while SVM showed lower but still substantial discriminative ability (18/25, 72%). XGB classification achieved an AUC of 0.80 ± 0.07, specificity of 0.80 ± 0.13, sensitivity of 0.81 ± 0.08, and F1 score of 0.86 ± 0.05, all comparable to RF and SVM classifiers (**Figure S9**, P > 0.05). The metrics indicate stable and moderate predictive ability of the overall water chemistry for distinguishing *Legionella* spp. occurrence. To minimize potential algorithm-specific bias when comparing predictors of abundance and occurrence, relative importance of individual variables from the XGB classification model was analyzed (**Figure 5G**). Among all predictors, pH emerged as the most influential variable for the occurrence of *Legionella* spp. Its high importance in both the regression (**Figure 5A**) and classification (**Figure 5G**) models likely reflects its close association with residual disinfectant chemistry and corresponding inactivation efficacy. In this study, chloraminated finished water exhibited significantly higher pH than chlorinated systems. Previous studies demonstrated that both free chlorine (pH 6–9) and monochloramine (pH 6–10) exhibited greater inactivation efficacy against *M. avium* as pH decreased within these ranges, reflecting increased oxidative efficacy of residual disinfectants under lower pH conditions^34,35^. Accordingly, pH may function as an integrated indicator of disinfectant regime and oxidative stress within the system. The prominence of pH in the classification model therefore suggests that disinfectant-associated oxidative pressure played a critical role in determining whether *Legionella* spp. could establish within DWDS. Its positive association with predicted abundance further indicates that higher pH conditions, associated with reduced oxidative pressure, may allow greater proliferation once colonized. In contrast, ammonia ranked among the least important predictors of *Legionella* spp. occurrence, suggesting that ammonia concentration alone was not a strong predictor of *Legionella* spp. occurrence but once present it could play an important role in its proliferation. Comparison of the XGB classification and regression models reveals complementary roles of pH and ammonia in shaping *Legionella* spp. persistence. Specifically, pH was the strongest predictor of occurrence, whereas ammonia was the strongest predictor of abundance. This distinction indicates stage-specific regulation within the overall chemical regime: pH could primarily regulate initial colonization by modulating disinfectant efficacy, while ammonia influences proliferation once *Legionella* spp. colonized, potentially through nitrification-mediated biofilm processes. Collectively, these findings highlight distinct but interconnected controls on *Legionella* spp. dynamics of overall water chemistry.

Regression models (RF, SVM, and XGB) exhibited poor predictive performance for *Mycobacterium* spp. abundance, with low or negative R^2^ (**Figure S10**, R² = −0.29 - −0.09), indicating that variation in abundance was not well explained by the measured water chemistry parameters. As a result, only classification models were further applied to evaluate how overall water chemistry conditions were associated with *Mycobacterium* spp. occurrence. The RF classifier yielded a mean AUC of 0.74 ± 0.09, statistically comparable to that of the SVM (0.72 ± 0.08) and XGB (0.70 ± 0.10; P > 0.05; **Figure S11**). Similarly, RF achieved a specificity of 0.75 ± 0.19, sensitivity of 0.74 ± 0.16, and F1 score of 0.79 ± 0.09, with no significant differences relative to the SVM and XGB classifiers (P > 0.05). These results indicate all three classifiers achieved comparable discriminatory performance; however, RF exhibited a higher proportion of statistically significant runs (**Figure S12**, 76%) across repeated cross-validation relative to SVM (36%) and XGB (48%), suggesting greater robustness under resampling. Accordingly, permutation-based variable importance from the RF classifier was used to identify key predictors (**Figure 5H**). Nitrate (16.3% ± 5.9%), sulfate (14.2% ± 7.5%), and pH (15.5% ± 6.0%) as the highest-ranking predictors, whereas ammonia ranked among the least influential variables. The relatively high importance of nitrate and sulfate, consistent with their contributions in the *Legionella* spp. classification models, suggests a broader role of redox gradients in shaping occurrence patterns. These results indicate that redox-associated processes, mediated by alternative electron acceptors, such as nitrate and sulfate, influence both microorganisms’ dynamics within DWDS. Overall, these findings indicate that both *Legionella* spp. and *Mycobacterium* spp. are shaped by the integrated water chemistry, with stage-specific effects and parameter-specific nonlinear response.

### Environmental Implications

This study demonstrates that integrated water chemistry, together with nonlinear, parameter-specific responses, are associated with *Mycobacterium* spp. and *Legionella* spp. occurrence and abundance in drinking water. These findings indicate that effective control should emphasize integrated water chemistry management in DWDS. Maintaining disinfectant-associated pH is critical for preventing their colonization, while controlling ammonia-driven nitrification in chloraminated systems is essential to limit their proliferation. The nonlinear response of DOC highlights the importance of identifying critical DOC ranges that minimize proliferation under specific integrated water chemical context. The importance of nitrate and sulfate further suggests that redox conditions should be incorporated into operational and monitoring strategies. Collectively, this work supports a transition from isolated parameter control toward integrated chemical regime management that simultaneously consider residual disinfectant, ammonia-driven nitrification, and redox conditions to regulate them in drinking water.

### Notes

The authors declare no competing financial interest.

## Data Availability

All data produced in the present study are available upon reasonable request to the authors

## Acknowledgments

This work was supported by the U.S. Environmental Protection Agency National Priorities Program (Grant No. 8406060). We also thank the participating water utility personnel for their assistance in this study.

## Notes

### Competing Interest Statement

The authors have declared no competing interest.

### Funding Statement

This study was funded by U.S. Environmental Protection Agency National Priorities Program

## Reference

(1) Johansen, M. D.; Herrmann, J.-L.; Kremer, L. Non-tuberculous mycobacteria and the rise of Mycobacterium abscessus. Nature Reviews Microbiology 2020, 18 (7), 392–407. DOI: 10.1038/s41579-020-0331-1.

(2) Scholar, E. Diseases Caused by Atypical Mycobacterium Species. In xPharm: The Comprehensive Pharmacology Reference, Enna, S. J., Bylund, D. B. Eds.; Elsevier, 2007; pp 1–4.

(3) Yang, J.; Hu, Y.; Zhang, Y.; Zhou, S.; Meng, D.; Xia, S.; Wang, H. Deciphering the diversity and assemblage mechanisms of nontuberculous mycobacteria community in four drinking water distribution systems with different disinfectants. Science of the Total Environment 2024, 907, 168176. DOI: 10.1016/j.scitotenv.2023.168176.

(4) Ditommaso, S.; Giacomuzzi, M.; Memoli, G.; Garlasco, J.; Zotti, C. M. Comparison of BCYEα+AB agar and MWY agar for detection and enumeration of Legionella spp. in hospital water samples. BMC Microbiology 2021, 21 (1), 48. DOI: 10.1186/s12866-021-02109-1.

(5) Zhong, Y.; Shen, L.; Zhou, Y.; Sun, Y.; Fu, X.; Huang, H. The Global Burden and Trends of Legionella spp. Infection-Associated Diseases from 1990 to 2021: An Observational Study. J Epidemiol Glob Health 2025, 15 (1), 3. DOI: 10.1007/s44197-025-00342-9 From NLM.

(6) Collier, S. A.; Deng, L.; Adam, E. A.; Benedict, K. M.; Beshearse, E. M.; Blackstock, A. J.; Bruce, B. B.; Derado, G.; Edens, C.; Fullerton, K. E.;, et al. Estimate of Burden and Direct Healthcare Cost of Infectious Waterborne Disease in the United States. Emerging Infectious Diseases 2021, 27 (1), 140–149. DOI: 10.3201/eid2701.190676.

(7) Barskey, A. E.; Derado, G.; Edens, C. Rising Incidence of Legionnaires’ Disease and Associated Epidemiologic Patterns, United States, 1992-2018. Emerg Infect Dis 2022, 28 (3), 527–538. DOI: 10.3201/eid2803.211435 From NLM.

(8) Moffa, M. A.; Rock, C.; Galiatsatos, P.; Gamage, S. D.; Schwab, K. J.; Exum, N. G. Legionellosis on the rise: A scoping review of sporadic, community-acquired incidence in the United States. Epidemiol Infect 2023, 151, e133. DOI: 10.1017/s0950268823001206 From NLM.

(9) Shah, N. M.; Davidson, J. A.; Anderson, L. F.; Lalor, M. K.; Kim, J.; Thomas, H. L.; Lipman, M.; Abubakar, I. Pulmonary Mycobacterium avium-intracellulare is the main driver of the rise in non-tuberculous mycobacteria incidence in England, Wales and Northern Ireland, 2007–2012. BMC Infectious Diseases 2016, 16 (1), 195. DOI: 10.1186/s12879-016-1521-3.

(10) Donohue, M. J. Increasing nontuberculous mycobacteria reporting rates and species diversity identified in clinical laboratory reports. BMC Infect Dis 2018, 18 (1), 163. DOI: 10.1186/s12879-018-3043-7 From NLM.

(11) Holsinger, H.; Tucker, N.; Regli, S.; Studer, K.; Roberts, V. A.; Collier, S.; Hannapel, E.; Edens, C.; Yoder, J. S.; Rotert, K. Characterization of reported legionellosis outbreaks associated with buildings served by public drinking water systems: United States, 2001–2017. Journal of Water and Health 2022, 20 (4), 702–711. DOI: 10.2166/wh.2022.002 (acccessed 2/11/2026).

(12) Buchholz, U.; Jahn, H. J.; Brodhun, B.; Lehfeld, A.-S.; Lewandowsky, M. M.; Reber, F.; Adler, K.; Bochmann, J.; Förster, C.; Koch, M.; et al. Source attribution of community-acquired cases of Legionnaires’ disease–results from the German LeTriWa study; Berlin, 2016–2019. Plos One 2020, 15 (11), e0241724. DOI: 10.1371/journal.pone.0241724.

(13) Almonacid Garrido, M. C.; Villanueva-Suárez, M. J.; Montes Martín, M. J.; Garcia-Alonso, A.; Tenorio Sanz, M. D. Prevalence and distribution of Legionella in municipal drinking water supply systems in Madrid (Spain) and risk factors associated. Science of the Total Environment 2024, 954, 176655. DOI: 10.1016/j.scitotenv.2024.176655.

(14) Waak, M. B.; LaPara, T. M.; Hallé, C.; Hozalski, R. M. Nontuberculous Mycobacteria in Two Drinking Water Distribution Systems and the Role of Residual Disinfection. Environmental Science & Technology 2019, 53 (15), 8563–8573. DOI: 10.1021/acs.est.9b01945.

(15) Luo, T.; Xu, P.; Zhang, Y.; Porter, J. L.; Ghanem, M.; Liu, Q.; Jiang, Y.; Li, J.; Miao, Q.; Hu, B.;, et al. Population genomics provides insights into the evolution and adaptation to humans of the waterborne pathogen Mycobacterium kansasii. Nature Communications 2021, 12 (1), 2491. DOI: 10.1038/s41467-021-22760-6.

(16) Dowdell, K.; Haig, S.-J.; Caverly, L. J.; Shen, Y.; LiPuma, J. J.; Raskin, L. Nontuberculous mycobacteria in drinking water systems – the challenges of characterization and risk mitigation. Current Opinion in Biotechnology 2019, 57, 127–136. DOI: 10.1016/j.copbio.2019.03.010.

(17) Donohue, M. J.; Vesper, S.; Mistry, J.; Donohue, J. M. Impact of Chlorine and Chloramine on the Detection and Quantification of Legionella pneumophila and Mycobacterium Species. Appl Environ Microbiol 2019, 85 (24). DOI: 10.1128/aem.01942-19 From NLM.

(18) Xi, H.; Ross, K. E.; Hinds, J.; Molino, P. J.; Whiley, H. Efficacy of chlorine-based disinfectants to control Legionella within premise plumbing systems. Water Research 2024, 259, 121794. DOI: 10.1016/j.watres.2024.121794.

(19) Legislature, N. J. Requires DEP, DOH, owners or operators of certain public community water systems, and owners or operators of certain buildings or facilities to take certain actions to prevent and control cases of Legionnaires’ disease. 221st Leg., Reg. Sess. ed.; 2024.

(20) Rhoads, W. J.; Pruden, A.; Edwards, M. A. Interactive Effects of Corrosion, Copper, and Chloramines on Legionella and Mycobacteria in Hot Water Plumbing. Environmental Science & Technology 2017, 51 (12), 7065–7075. DOI: 10.1021/acs.est.6b05616.

(21) Marchesi, I.; Ferranti, G.; Mansi, A.; Marcelloni, A. M.; Proietto, A. R.; Saini, N.; Borella, P.; Bargellini, A. Control of Legionella Contamination and Risk of Corrosion in Hospital Water Networks following Various Disinfection Procedures. Applied and Environmental Microbiology 2016, 82 (10), 2959–2965. DOI: doi:10.1128/AEM.03873-15.

(22) Proctor, C. R.; Dai, D.; Edwards, M. A.; Pruden, A. Interactive effects of temperature, organic carbon, and pipe material on microbiota composition and Legionella pneumophila in hot water plumbing systems. Microbiome 2017, 5 (1), 130. DOI: 10.1186/s40168-017-0348-5.

(23) Li, H.; Li, S.; Tang, W.; Yang, Y.; Zhao, J.; Xia, S.; Zhang, W.; Wang, H. Influence of secondary water supply systems on microbial community structure and opportunistic pathogen gene markers. Water Research 2018, 136, 160–168. DOI: 10.1016/j.watres.2018.02.031.

(24) van der Wielen, P. W.; van der Kooij, D. Nontuberculous mycobacteria, fungi, and opportunistic pathogens in unchlorinated drinking water in The Netherlands. Appl Environ Microbiol 2013, 79 (3), 825–834. DOI: 10.1128/aem.02748-12 From NLM.

(25) Schwake, D. O.; Garner, E.; Strom, O. R.; Pruden, A.; Edwards, M. A. Legionella DNA Markers in Tap Water Coincident with a Spike in Legionnaires’ Disease in Flint, MI. Environmental Science & Technology Letters 2016, 3 (9), 311–315. DOI: 10.1021/acs.estlett.6b00192.

(26) D’Alessandro, D.; Fabiani, M.; Cerquetani, F.; Orsi, G. B. Trend of Legionella colonization in hospital water supply. Ann Ig 2015, 27 (2), 460–466. DOI: 10.7416/ai.2015.2032 From NLM.

(27) Falkinham, J. O, 3rd.; Norton, C. D.; LeChevallier, M. W. Factors influencing numbers of Mycobacterium avium, Mycobacterium intracellulare, and other Mycobacteria in drinking water distribution systems. Appl Environ Microbiol 2001, 67 (3), 1225–1231. DOI: 10.1128/aem.67.3.1225-1231.2001 From NLM.

(28) Torvinen, E.; Suomalainen, S.; Lehtola, M. J.; Miettinen, I. T.; Zacheus, O.; Paulin, L.; Katila, M. L.; Martikainen, P. J. Mycobacteria in water and loose deposits of drinking water distribution systems in Finland. Appl Environ Microbiol 2004, 70 (4), 1973–1981. DOI: 10.1128/aem.70.4.1973-1981.2004 From NLM.

(29) Falkinham, I. J. O. Surrounded by mycobacteria: nontuberculous mycobacteria in the human environment. Journal of Applied Microbiology 2009, 107 (2), 356–367. DOI: 10.1111/j.1365-2672.2009.04161.x (acccessed 2026/02/11).

(30) Wullings, B. A.; Bakker, G.; Kooij, D. v. d. Concentration and Diversity of Uncultured Legionella spp. in Two Unchlorinated Drinking Water Supplies with Different Concentrations of Natural Organic Matter. Applied and Environmental Microbiology 2011, 77 (2), 634–641. DOI: doi:10.1128/AEM.01215-10.

(31) Norton, C. D.; LeChevallier, M. W.; Falkinham, J. O. Survival of Mycobacterium avium in a model distribution system. Water Research 2004, 38 (6), 1457–1466. DOI: 10.1016/j.watres.2003.07.008.

(32) Duda, S.; Kandiah, S.; Stout, J. E.; Baron, J. L.; Yassin, M.; Fabrizio, M.; Ferrelli, J.; Hariri, R.; Wagener, M. M.; Goepfert, J.;, et al. Evaluation of a New Monochloramine Generation System for Controlling Legionella in Building Hot Water Systems. Infection Control & Hospital Epidemiology 2014, 35 (11), 1356–1363. DOI: 10.1086/678418 From Cambridge University Press Cambridge Core.

(33) Leoni, E.; De Luca, G.; Legnani, P. P.; Sacchetti, R.; Stampi, S.; Zanetti, F. Legionella waterline colonization: detection of Legionella species in domestic, hotel and hospital hot water systems. J Appl Microbiol 2005, 98 (2), 373–379. DOI: 10.1111/j.1365-2672.2004.02458.x From NLM Medline.

(34) Luh, J.; Mariñas, B. J. Inactivation of Mycobacterium avium with Free Chlorine. Environmental Science & Technology 2007, 41 (14), 5096–5102. DOI: 10.1021/es0630801.

(35) Luh, J.; Tong, N.; Raskin, L.; Mariñas, B. J. Inactivation of Mycobacterium avium with Monochloramine. Environmental Science & Technology 2008, 42 (21), 8051–8056. DOI: 10.1021/es801133q.

(36) Vicuña-Reyes, J. P.; Luh, J.; Mariñas, B. J. Inactivation of Mycobacterium avium with chlorine dioxide. Water Research 2008, 42 (6), 1531–1538. DOI: 10.1016/j.watres.2007.10.035.

(37) Pang, H.; Ben, Y.; Cao, Y.; Qu, S.; Hu, C. Time series-based machine learning for forecasting multivariate water quality in full-scale drinking water treatment with various reagent dosages. Water Research 2025, 268, 122777. DOI: 10.1016/j.watres.2024.122777.

(38) Wang, Y.-Q.; Wang, H.-C.; Xiao, Z.-J.; Bu, L.-J.; Li, J.; Feng, X.-C.; Liang, B.; Liu, W.-Z.; Sun, F.-Y.; Zhou, S.-Q.;, et al. Machine learning strategy secures urban smart drinking water treatment plant through incremental advances. Water Research 2025, 280, 123541. DOI: 10.1016/j.watres.2025.123541.

(39) Perelman, L.; Arad, J.; Housh, M.; Ostfeld, A. Event Detection in Water Distribution Systems from Multivariate Water Quality Time Series. Environmental Science & Technology 2012, 46 (15), 8212–8219. DOI: 10.1021/es3014024.

(40) Oliker, N.; Ostfeld, A. Comparison of two multivariate classification models for contamination event detection in water quality time series. Journal of Water Supply: Research and Technology-Aqua 2014, 64 (5), 558–566. DOI: 10.2166/aqua.2014.033 (acccessed 2/12/2026).

(41) Brown, M. G. L.; Peterson, M. G.; Tezaur, I. K.; Peterson, K. J.; Bull, D. L. Random forest regression feature importance for climate impact pathway detection. Journal of Computational and Applied Mathematics 2025, 464, 116479. DOI: 10.1016/j.cam.2024.116479.

(42) Wang, F.; Wang, Y.; Zhang, K.; Hu, M.; Weng, Q.; Zhang, H. Spatial heterogeneity modeling of water quality based on random forest regression and model interpretation. Environmental Research 2021, 202, 111660. DOI: 10.1016/j.envres.2021.111660.

(43) Brester, C.; Ryzhikov, I.; Siponen, S.; Jayaprakash, B.; Ikonen, J.; Pitkänen, T.; Miettinen, I. T.; Torvinen, E.; Kolehmainen, M. Potential and limitations of a pilot-scale drinking water distribution system for bacterial community predictive modelling. Science of the Total Environment 2020, 717, 137249. DOI: 10.1016/j.scitotenv.2020.137249.

(44) Archer, K. J.; Kimes, R. V. Empirical characterization of random forest variable importance measures. Computational Statistics & Data Analysis 2008, 52 (4), 2249–2260. DOI: 10.1016/j.csda.2007.08.015.

(45) Luo, Z. N.; He, H.; Zhang, T. Y.; Wei, X. L.; Dong, Z. Y.; Xu, M. Y.; Zhao, H. X.; Zheng, Z. X.; Pan, R. J.; Hu, C. Y.;, et al. Enhanced iodinated disinfection byproducts formation in iodide/ iodate-containing water undergoing UV-chloramine sequential disinfection: Machine learning-aided identification of reaction mechanisms. Water Research 2025, 272. DOI: 10.1016/j.watres.2024.122975.

(46) Ransom, K. M.; Nolan, B. T.; Stackelberg, P. E.; Belitz, K.; Fram, M. S. Machine learning predictions of nitrate in groundwater used for drinking supply in the conterminous United States. Science of the Total Environment 2022, 807. DOI: 10.1016/j.scitotenv.2021.151065.

(47) Park, J.; Lee, W. H.; Kim, K. T.; Park, C. Y.; Lee, S.; Heo, T. Y. Interpretation of ensemble learning to predict water quality using explainable artificial intelligence. Science of the Total Environment 2022, 832. DOI: 10.1016/j.scitotenv.2022.155070.

(48) Chatterjee, D.; Ghosh, P.; Banerjee, A.; Das, S. S. Optimizing machine learning for water safety: A comparative analysis with dimensionality reduction and classifier performance in potability prediction. PLOS Water 2024, 3 (8), e0000259. DOI: 10.1371/journal.pwat.0000259.

(49) Li, H.; Zhou, B.; Xu, X.; Huo, R.; Zhou, T.; Dong, X.; Ye, C.; Li, T.; Xie, L.; Pang, W. The insightful water quality analysis and predictive model establishment via machine learning in dual-source drinking water distribution system. Environmental Research 2024, 250, 118474. DOI: 10.1016/j.envres.2024.118474.

(50) Das, A. Surface water quality evaluation impacting drinking water sources and sanitation using water quality index, multivariate techniques, and interpretable machine learning models in Mahanadi River, Odisha (India). Environmental Geochemistry and Health 2025, 47 (11), 497. DOI: 10.1007/s10653-025-02806-0.

(51) Apley, D. W.; Zhu, J. Visualizing the Effects of Predictor Variables in Black Box Supervised Learning Models. Journal of the Royal Statistical Society Series B: Statistical Methodology 2020, 82 (4), 1059–1086. DOI: 10.1111/rssb.12377 (acccessed 12/7/2025).

(52) DiLoreto, S.; He, H.; Yang, J.; Milne, P.; Li, J.; Impellitteri, C. A.; Stubbins, A.; Pinto, A.; Huang, C. H. Identification of Factors Influencing Variability in Disinfection Byproducts and Their Toxicity in Chlorinated and Chloraminated Drinking Water Distribution Systems across the United States. Environ Sci Technol 2026, 60 (1), 1241–1252. DOI: 10.1021/acs.est.5c12121 From NLM Medline.

(53) Vosloo, S.; Sevillano, M.; Pinto, A. Modified DNeasy PowerWater Kit® protocol for DNA extractions from drinking water samples. protocolsio. 2019.

(54) He, H.; DiLoreto, S.; Yang, J.; Milne, P.; Impellitteri, C. A.; Stubbins, A.; Pieper, K.; Graham, K.; Huang, C.-H.; Pinto, A. <em>Legionella</em> and <em>Mycobacterium</em> populations exhibit geographic structuring across and within drinking water systems. bioRxiv 2026, 2026.2001.2013.699378. DOI: 10.64898/2026.01.13.699378.

(55) Nazarian, E. J.; Bopp, D. J.; Saylors, A.; Limberger, R. J.; Musser, K. A. Design and implementation of a protocol for the detection of Legionella in clinical and environmental samples. Diagnostic Microbiology and Infectious Disease 2008, 62 (2), 125–132. DOI: 10.1016/j.diagmicrobio.2008.05.004.

(56) Radomski, N.; Lucas Françoise, S.; Moilleron, R.; Cambau, E.; Haenn, S.; Moulin, L. Development of a Real-Time qPCR Method for Detection and Enumeration of Mycobacterium spp. in Surface Water. Applied and Environmental Microbiology 2010, 76 (21), 7348–7351. DOI: 10.1128/AEM.00942-10 (acccessed 2026/02/16).

(57) García-Quintanilla, A.; González-Martín, J.; Tudó, G.; Espasa, M.; Anta, M. T. J. d. Simultaneous Identification of Mycobacterium Genus and Mycobacterium tuberculosis Complex in Clinical Samples by 5&#x2032;-Exonuclease Fluorogenic PCR. Journal of Clinical Microbiology 2002, 40 (12), 4646–4651. DOI: doi:10.1128/jcm.40.12.4646-4651.2002.

(58) Wang, H.; Edwards, M.; Falkinham Joseph, O.; Pruden, A. Molecular Survey of the Occurrence of Legionella spp., Mycobacterium spp., Pseudomonas aeruginosa, and Amoeba Hosts in Two Chloraminated Drinking Water Distribution Systems. Applied and Environmental Microbiology 2012, 78 (17), 6285–6294. DOI: 10.1128/AEM.01492-12 (acccessed 2026/02/16).

(59) Armbruster, D. A.; Pry, T. Limit of blank, limit of detection and limit of quantitation. Clin Biochem Rev 2008, 29 *Suppl 1* (Suppl 1), S49-52. From NLM.

(60) R: A Language and Environment for Statistical Computing; R Foundation for Statistical Computing: Vienna, Austria, 2025. https://www.R-project.org/ (accessed.

(61) Yu, W.; Yang, J.; Cai, X.; Xia, S.; Wang, H. Unraveling the dynamics and growth potential of non-tuberculous mycobacteria along water transportation in chloraminated secondary water supply systems. Environmental Pollution 2025, 383, 126812. DOI: 10.1016/j.envpol.2025.126812.

(62) Pfaller, S.; King, D.; Mistry, J. H.; Alexander, M.; Abulikemu, G.; Pressman, J. G.; Wahman, D. G.; Donohue, M. J. Chloramine Concentrations within Distribution Systems and Their Effect on Heterotrophic Bacteria, Mycobacterial Species, and Disinfection Byproducts. Water Research 2021, 205, 117689. DOI: 10.1016/j.watres.2021.117689.

(63) Hull, N. M.; Holinger, E. P.; Ross, K. A.; Robertson, C. E.; Harris, J. K.; Stevens, M. J.; Pace, N. R. Longitudinal and Source-to-Tap New Orleans, LA, U.S.A. Drinking Water Microbiology. Environmental Science & Technology 2017, 51 (8), 4220–4229. DOI: 10.1021/acs.est.6b06064.

(64) Gan, Y.; Kurisu, F.; Simazaki, D.; Yoshida, M.; Fukano, H.; Komine, T.; Nagashima, H.; Hoshino, Y.; Kasuga, I. Unveiling significant regrowth and potential risk of nontuberculous mycobacteria in hospital water supply system. Water Research 2025, 275, 123188. DOI: 10.1016/j.watres.2025.123188.

(65) Shen, Y.; Huang, C.; Lin, J.; Wu, W.; Ashbolt, N. J.; Liu, W.-T.; Nguyen, T. H. Effect of Disinfectant Exposure on Legionella pneumophila Associated with Simulated Drinking Water Biofilms: Release, Inactivation, and Infectivity. Environmental Science & Technology 2017, 51 (4), 2087–2095. DOI: 10.1021/acs.est.6b04754.

(66) Shen, Y.; Monroy, G. L.; Derlon, N.; Janjaroen, D.; Huang, C.; Morgenroth, E.; Boppart, S. A.; Ashbolt, N. J.; Liu, W.-T.; Nguyen, T. H. Role of Biofilm Roughness and Hydrodynamic Conditions in Legionella pneumophila Adhesion to and Detachment from Simulated Drinking Water Biofilms. Environmental Science & Technology 2015, 49 (7), 4274–4282. DOI: 10.1021/es505842v.

(67) Douterelo, I.; Husband, S.; Loza, V.; Boxall, J. Dynamics of Biofilm Regrowth in Drinking Water Distribution Systems. Applied and Environmental Microbiology 2016, 82 (14), 4155–4168. DOI: 10.1128/AEM.00109-16 (acccessed 2026/02/12).

(68) Armbruster, C. R.; Forster, T. S.; Donlan, R. M.; O’Connell, H. A.; Shams, A. M.; Williams, M. M. A biofilm model developed to investigate survival and disinfection of Mycobacterium mucogenicum in potable water. Biofouling 2012, 28 (10), 1129–1139. DOI: 10.1080/08927014.2012.735231.

(69) Wang, H.; Masters, S.; Hong, Y.; Stallings, J.; Falkinham, J. O., III; Edwards, M. A.; Pruden, A. Effect of Disinfectant, Water Age, and Pipe Material on Occurrence and Persistence of Legionella, mycobacteria, Pseudomonas aeruginosa, and Two Amoebas. Environmental Science & Technology 2012, 46 (21), 11566–11574. DOI: 10.1021/es303212a.

(70) Pressman, J. G.; Lee, W. H.; Bishop, P. L.; Wahman, D. G. Effect of free ammonia concentration on monochloramine penetration within a nitrifying biofilm and its effect on activity, viability, and recovery. Water Research 2012, 46 (3), 882–894. DOI: 10.1016/j.watres.2011.11.071.

(71) Keshvardoust, P.; Huron, V. A. A.; Clemson, M.; Constancias, F.; Barraud, N.; Rice, S. A. Biofilm formation inhibition and dispersal of multi-species communities containing ammonia-oxidising bacteria. npj Biofilms and Microbiomes 2019, 5 (1), 22. DOI: 10.1038/s41522-019-0095-4.

(72) Xue, Z.; Sendamangalam, V. R.; Gruden, C. L.; Seo, Y. Multiple Roles of Extracellular Polymeric Substances on Resistance of Biofilm and Detached Clusters. Environmental Science & Technology 2012, 46 (24), 13212–13219. DOI: 10.1021/es3031165.

(73) Donohue, M. J.; Vesper, S.; Mistry, J.; Donohue, J. M. Impact of Chlorine and Chloramine on the Detection and Quantification of Legionella pneumophila and Mycobacterium Species. Applied and Environmental Microbiology 2019, 85 (24), e01942–01919. DOI: doi:10.1128/AEM.01942-19.

(74) Muñoz Egea, M. C.; Ji, P.; Pruden, A.; Falkinham Iii, J. O. Inhibition of Adherence of Mycobacterium avium to Plumbing Surface Biofilms of Methylobacterium spp. Pathogens 2017, 6 (3). DOI: 10.3390/pathogens6030042 From NLM.

(75) Falkinham, J. O 3rd.; Williams, M. D.; Kwait, R.; Lande, L. Methylobacterium spp. as an indicator for the presence or absence of Mycobacterium spp. Int J Mycobacteriol 2016, 5 (2), 240–243. DOI: 10.1016/j.ijmyco.2016.03.001 From NLM.

(76) Drancourt, M. Looking in amoebae as a source of mycobacteria. Microbial

(77) Delafont, V.; Mougari, F.; Cambau, E.; Joyeux, M.; Bouchon, D.; Héchard, Y.; Moulin, L. First Evidence of Amoebae–Mycobacteria Association in Drinking Water Network. Environmental Science & Technology 2014, 48 (20), 11872–11882. DOI: 10.1021/es5036255.

(78) Thomas, V.; McDonnell, G.; Denyer, S. P.; Maillard, J. Y. Free-living amoebae and their intracellular pathogenic microorganisms: risks for water quality. FEMS Microbiol Rev 2010, 34 (3), 231–259. DOI: 10.1111/j.1574-6976.2009.00190.x From NLM.

(79) Dietersdorfer, E.; Kirschner, A.; Schrammel, B.; Ohradanova-Repic, A.; Stockinger, H.; Sommer, R.; Walochnik, J.; Cervero-Aragó, S. Starved viable but non-culturable (VBNC) Legionella strains can infect and replicate in amoebae and human macrophages. Water Research 2018, 141, 428–438. DOI: 10.1016/j.watres.2018.01.058.

(80) Shaheen, M.; Scott, C.; Ashbolt, N. J. Long-term persistence of infectious Legionella with free-living amoebae in drinking water biofilms. International Journal of Hygiene and Environmental Health 2019, 222 (4), 678–686. DOI: 10.1016/j.ijheh.2019.04.007.

(81) König, L.; Wentrup, C.; Schulz, F.; Wascher, F.; Escola, S.; Swanson Michele, S.; Buchrieser, C.; Horn, M. Symbiont-Mediated Defense against Legionella pneumophila in Amoebae. Mbio 2019, 10 (3),. DOI: 10.1128/mbio.00333-19 (acccessed 2026/02/14).

(82) Steed, K. A.; Falkinham, J. O., 3rd. Effect of growth in biofilms on chlorine susceptibility of Mycobacterium avium and Mycobacterium intracellulare. Appl Environ Microbiol 2006, 72 (6), 4007–4011. DOI: 10.1128/aem.02573-05 From NLM.

(83) Cazals, M.; Bédard, E.; Faucher, S. P.; Prévost, M. Factors Affecting the Dynamics of Legionella pneumophila, Nontuberculous Mycobacteria, and Their Host Vermamoeba vermiformis in Premise Plumbing. Acs Es&T Water 2023, 3 (12), 3874–3883. DOI: 10.1021/acsestwater.3c00288.

(84) Mathys, W.; Stanke, J.; Harmuth, M.; Junge-Mathys, E. Occurrence of Legionella in hot water systems of single-family residences in suburbs of two German cities with special reference to solar and district heating. International Journal of Hygiene and Environmental Health 2008, 211 (1), 179–185. DOI: 10.1016/j.ijheh.2007.02.004.

(85) Haig, S.-J.; Kotlarz, N.; LiPuma, J. J.; Raskin, L. A High-Throughput Approach for Identification of Nontuberculous Mycobacteria in Drinking Water Reveals Relationship between Water Age and Mycobacterium avium. Mbio 2018, 9 (1),. DOI: doi:10.1128/mbio.02354-17.

(86) Ley, C. J.; Proctor, C. R.; Singh, G.; Ra, K.; Noh, Y.; Odimayomi, T.; Salehi, M.; Julien, R.; Mitchell, J.; Nejadhashemi, A. P.;, et al. Drinking water microbiology in a water-efficient building: stagnation, seasonality, and physicochemical effects on opportunistic pathogen and total bacteria proliferation. Environmental Science: Water Research & Technology 2020, 6 (10), 2902–2913. DOI: 10.1039/d0ew00334d.

(87) Cunliffe, D. A. Bacterial nitrification in chloraminated water supplies. Applied and Environmental Microbiology 1991, 57 (11), 3399–3402. DOI: 10.1128/aem.57.11.3399-3402.1991 (acccessed 2026/02/14).

(88) Woolschlager, J.; Rittmann, B.; Piriou, P.; Kiene, L.; Schwartz, B. Using a comprehensive model to identify the major mechanisms of chloramine decay in distribution systems. Water science and technology: water supply 2001, 1 (4), 103–110.

(89) Wolfe, R. L. III;, E. G. M. Davis, M. K. Barrett, S. E. Biological nitrification in covered reservoirs containing chloraminated water. Journal-American Water Works Association 1988, 80 (9), 109–114.

(90) Masters, S.; Wang, H.; Pruden, A.; Edwards, M. A. Redox gradients in distribution systems influence water quality, corrosion, and microbial ecology. Water Res 2015, 68, 140–149. DOI: 10.1016/j.watres.2014.09.048 From NLM Medline.

(91) Potgieter Sarah, C.; Dai, Z.; Venter Stephanus, N.; Sigudu, M.; Pinto Ameet, J. Microbial Nitrogen Metabolism in Chloraminated Drinking Water Reservoirs. Msphere 2020, 5 (2),. DOI: 10.1128/msphere.00274-20 (acccessed 2026/02/14).

(92) Lytle, D. A.; Gerke, T. L.; Maynard, D. J. B. Effect of bacterial sulfate reduction on iron-corrosion scales. Journal AWWA 2005, 97 (10), 109–120. DOI: 10.1002/j.1551-8833.2005.tb07500.x (acccessed 2026/02/14).

(93) Gomez-Smith, C. K.; LaPara, T. M.; Hozalski, R. M. Sulfate Reducing Bacteria and Mycobacteria Dominate the Biofilm Communities in a Chloraminated Drinking Water Distribution System. Environmental Science & Technology 2015, 49 (14), 8432–8440. DOI: 10.1021/acs.est.5b00555.

(94) Javaherdashti, R. Impact of sulphate-reducing bacteria on the performance of engineering materials. Appl Microbiol Biotechnol 2011, 91 (6), 1507–1517. DOI: 10.1007/s00253-011-3455-4 From NLM.

(95) Seth, A. D.; Edyvean, R. G. J. The function of sulfate-reducing bacteria in corrosion of potable water mains. International Biodeterioration & Biodegradation 2006, 58 (3), 108–111. DOI: 10.1016/j.ibiod.2006.10.005.

(96) Yang, F.; Shi, B.; Gu, J.; Wang, D.; Yang, M. Morphological and physicochemical characteristics of iron corrosion scales formed under different water source histories in a drinking water distribution system. Water Research 2012, 46 (16), 5423–5433. DOI: 10.1016/j.watres.2012.07.031.

(97) Morton, S. C.; Zhang, Y.; Edwards, M. A. Implications of nutrient release from iron metal for microbial regrowth in water distribution systems. Water Research 2005, 39 (13), 2883–2892. DOI: 10.1016/j.watres.2005.05.024.

