## supplemental figures for "Interpretable Machine Learning Reveals Integrated Water Chemistry and Parameter-Specific Nonlinear Responses Shaping *Legionella* spp. and *Mycobacterium* spp. in Drinking Water"

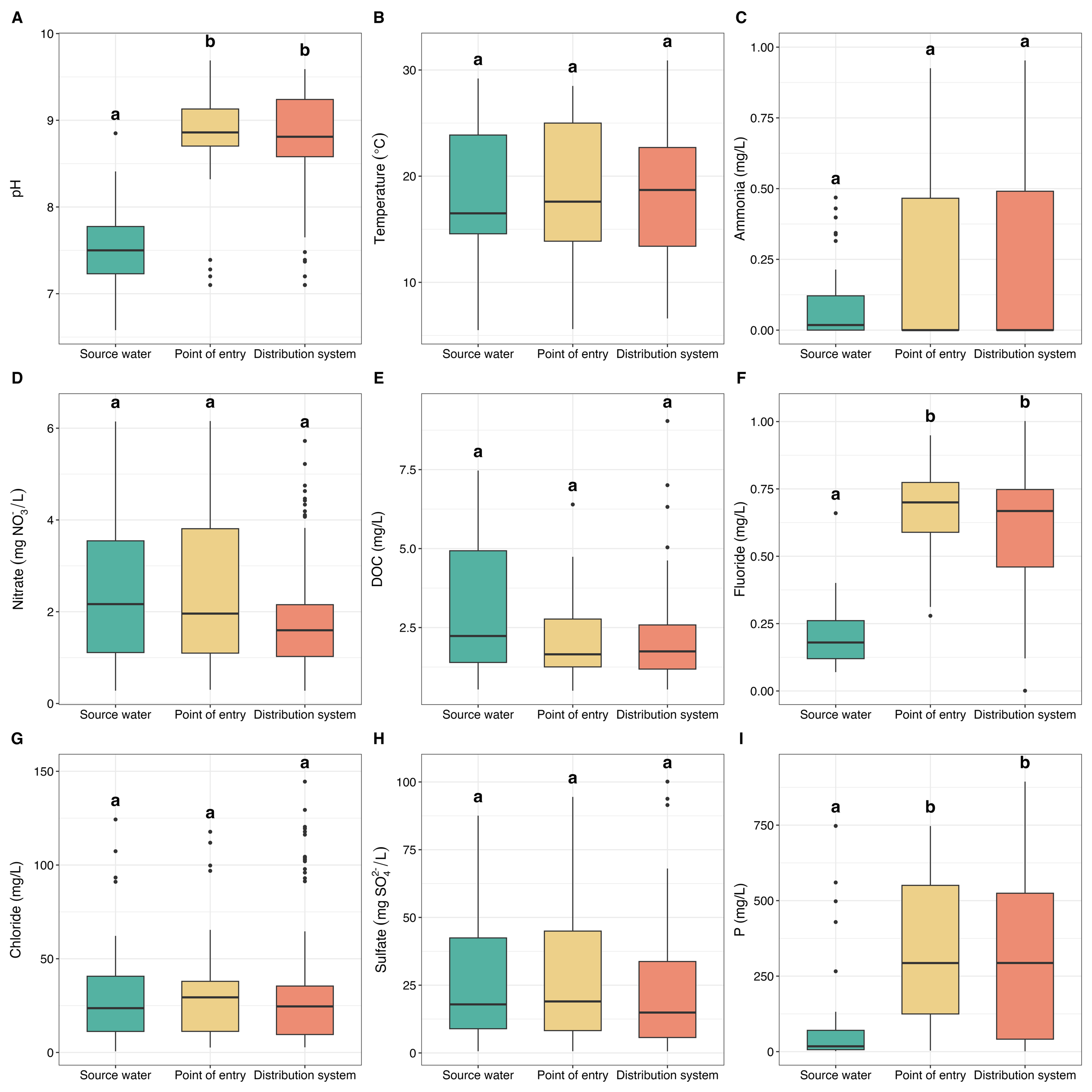

**Figure S1.** Water chemistry dynamics across source water, point-of-entry, and drinking water distribution system samples. (A) pH, (B) temperature, (C) ammonia, (D) nitrate, (E) DOC, (F) fluoride, (G) chloride, (H) sulfate, and (I) phosphorus. Different letters above boxplots denote statistically significant differences among sample types (P *<* 0.05).

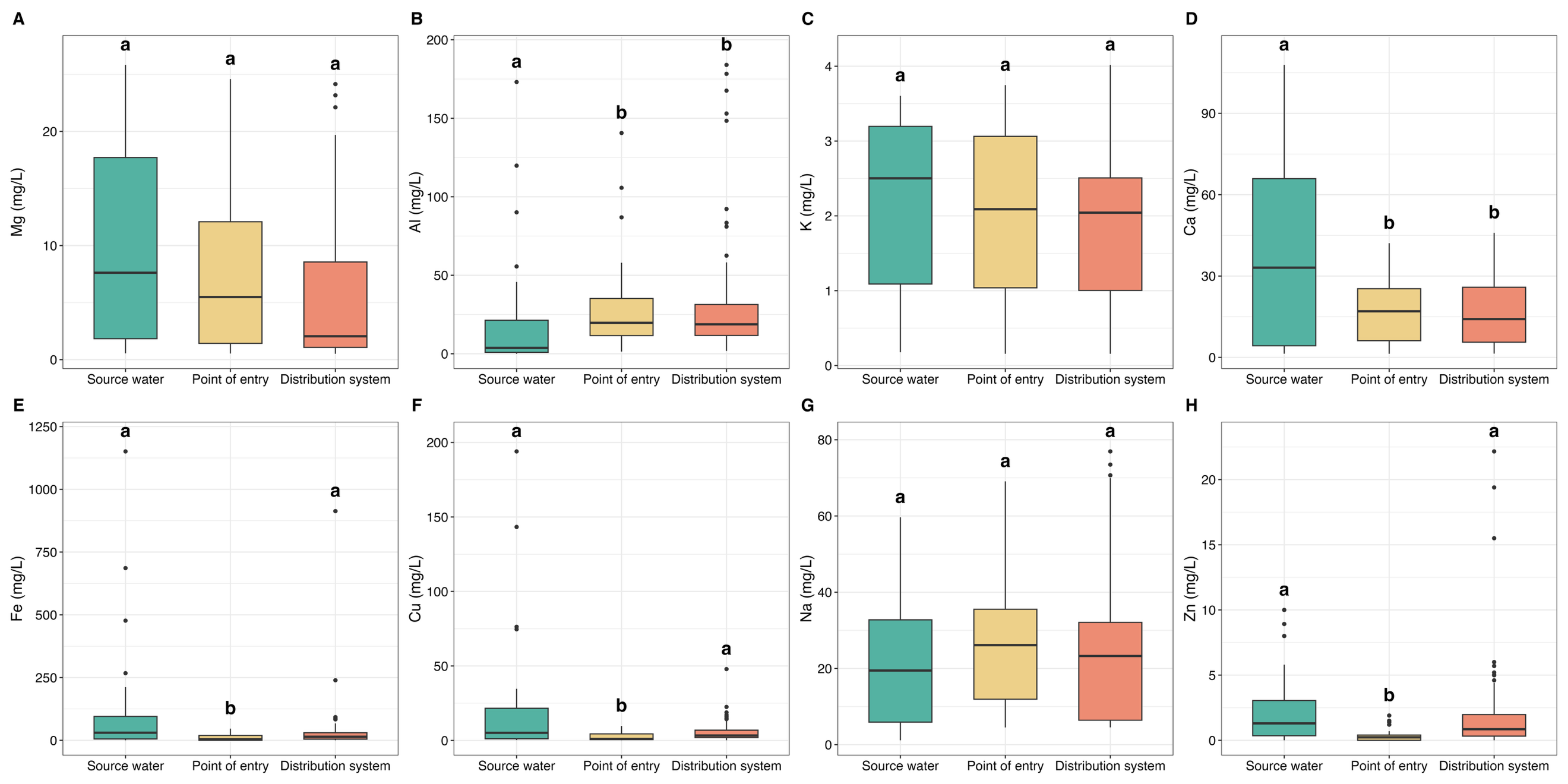

**Figure S2.** Water chemistry dynamics across source water, point-of-entry, and drinking water distribution system samples. (A) Mg, (B) Al, (C) K, (D) Ca, (E) Fe, (F) Cu, (G) Na, and (H) Zn. Different letters above boxplots denote statistically significant differences among sample types (P *<* 0.05).

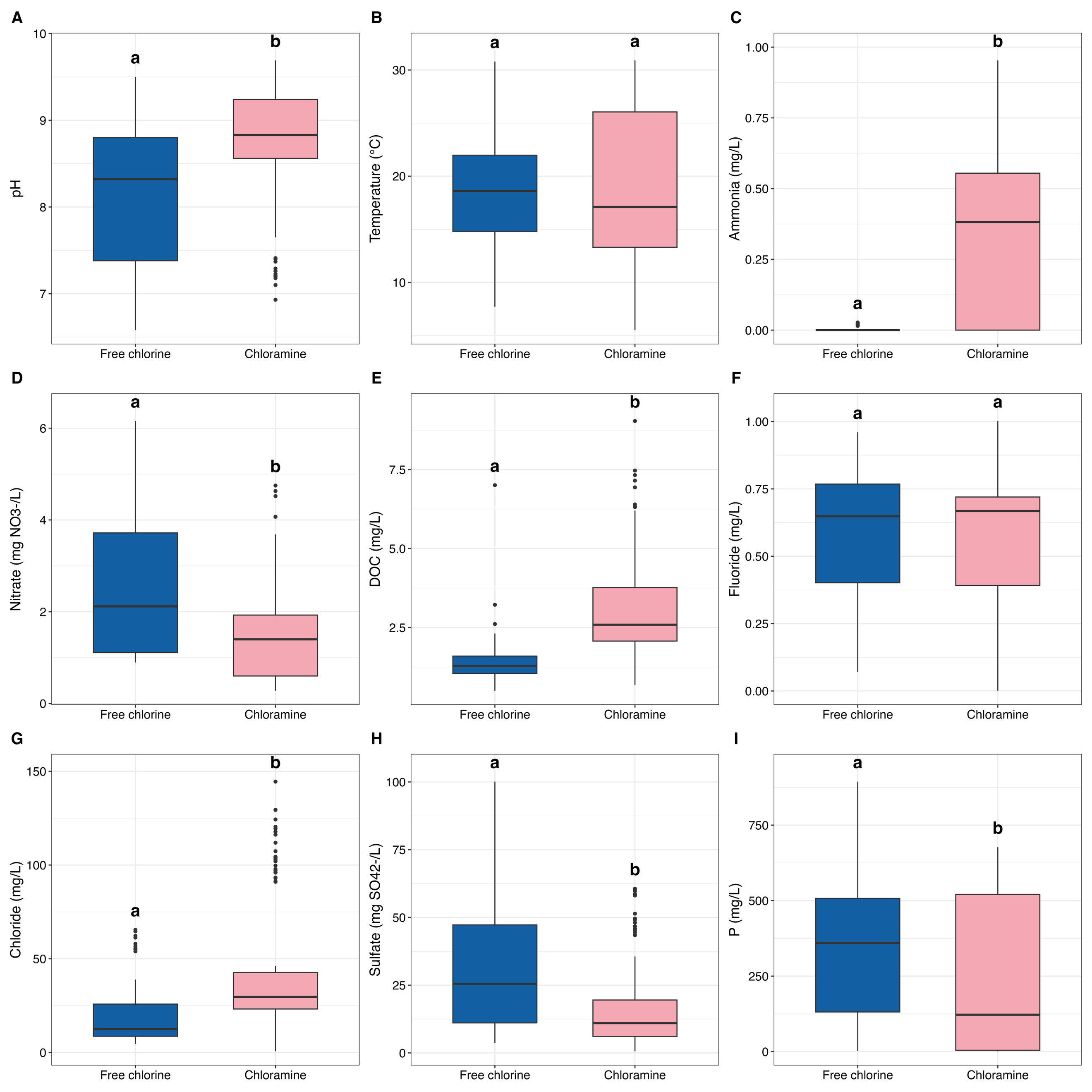

**Figure S3. Differences in water chemistry across free chlorine and chloramine drinking water distribution systems. (A) pH, (B) temperature, (C) ammonia, (D) nitrate, (E) DOC, (F) fluoride, (G) chloride, (H) sulfate, and (I) phosphorus. Different letters above boxplots denote statistically significant differences between disinfectant types (P < 0.05).**

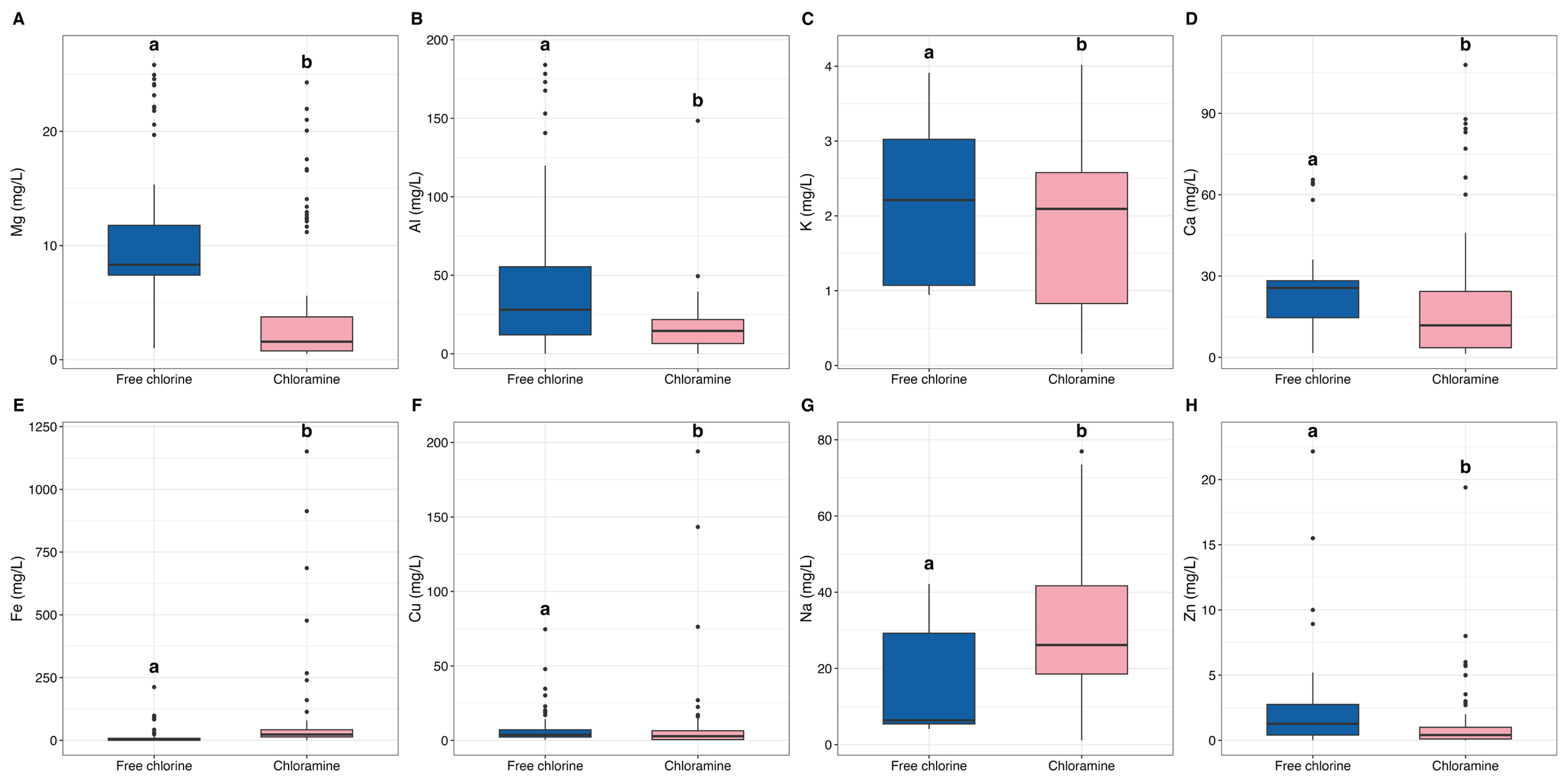

**Figure S4. Concentrations of (A) Mg, (B) Al, (C) K, (D) Ca, (E) Fe, (F) Cu, (G) Na, and (H) Zn in free chlorine and chloramine drinking water systems. Different letters above boxplots denote statistically significant differences between disinfectant types (P < 0.05).**

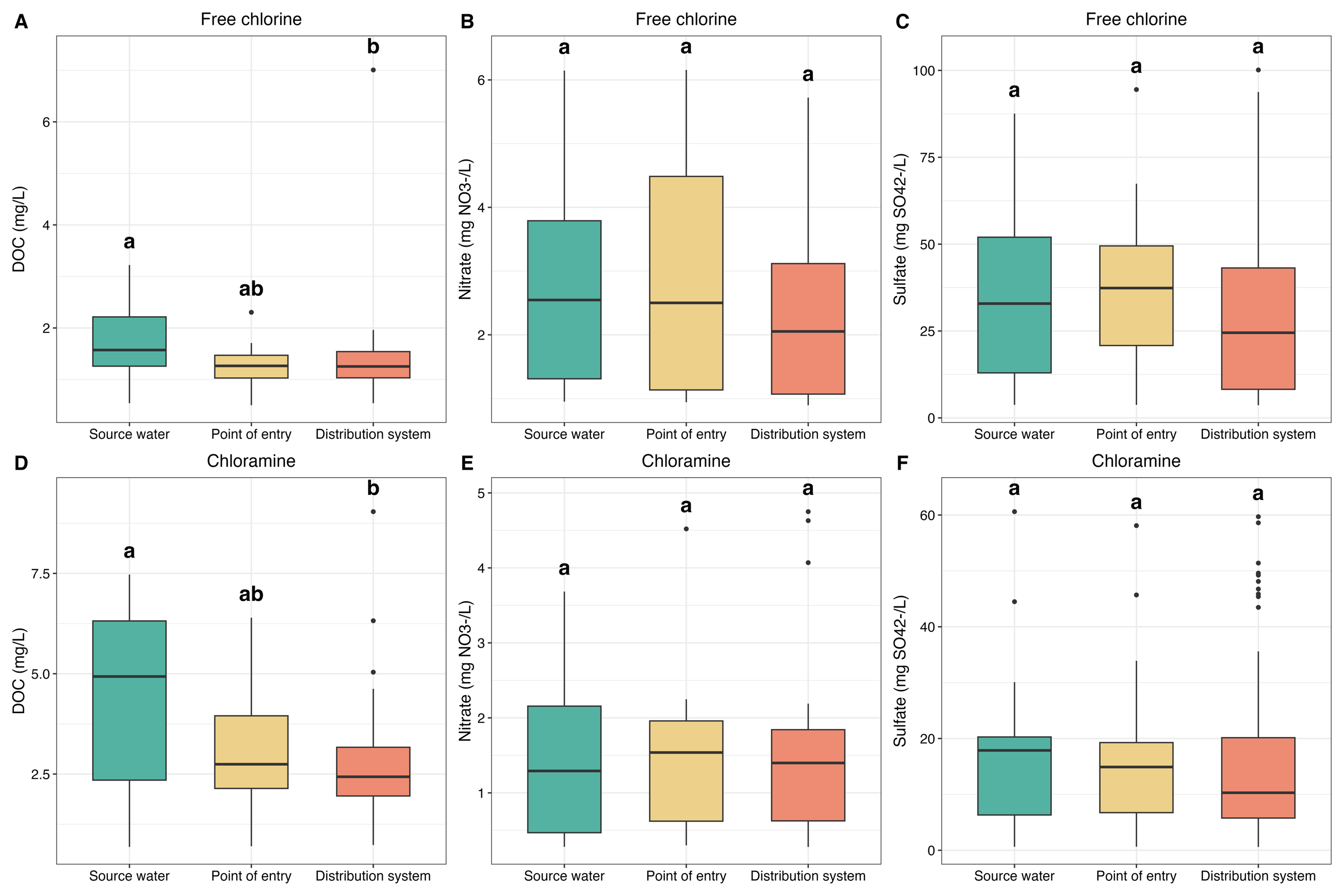

**Figure S5. Concentrations of DOC, nitrate, and sulfate across source water, point-of-entry, and distribution system samples within free chlorine and chloramine drinking water systems. The upper panel shows free chlorine systems, and the lower panel shows chloramine systems. Different letters above boxplots denote statistically significant differences among sample types within each disinfectant regime (P < 0.05).**

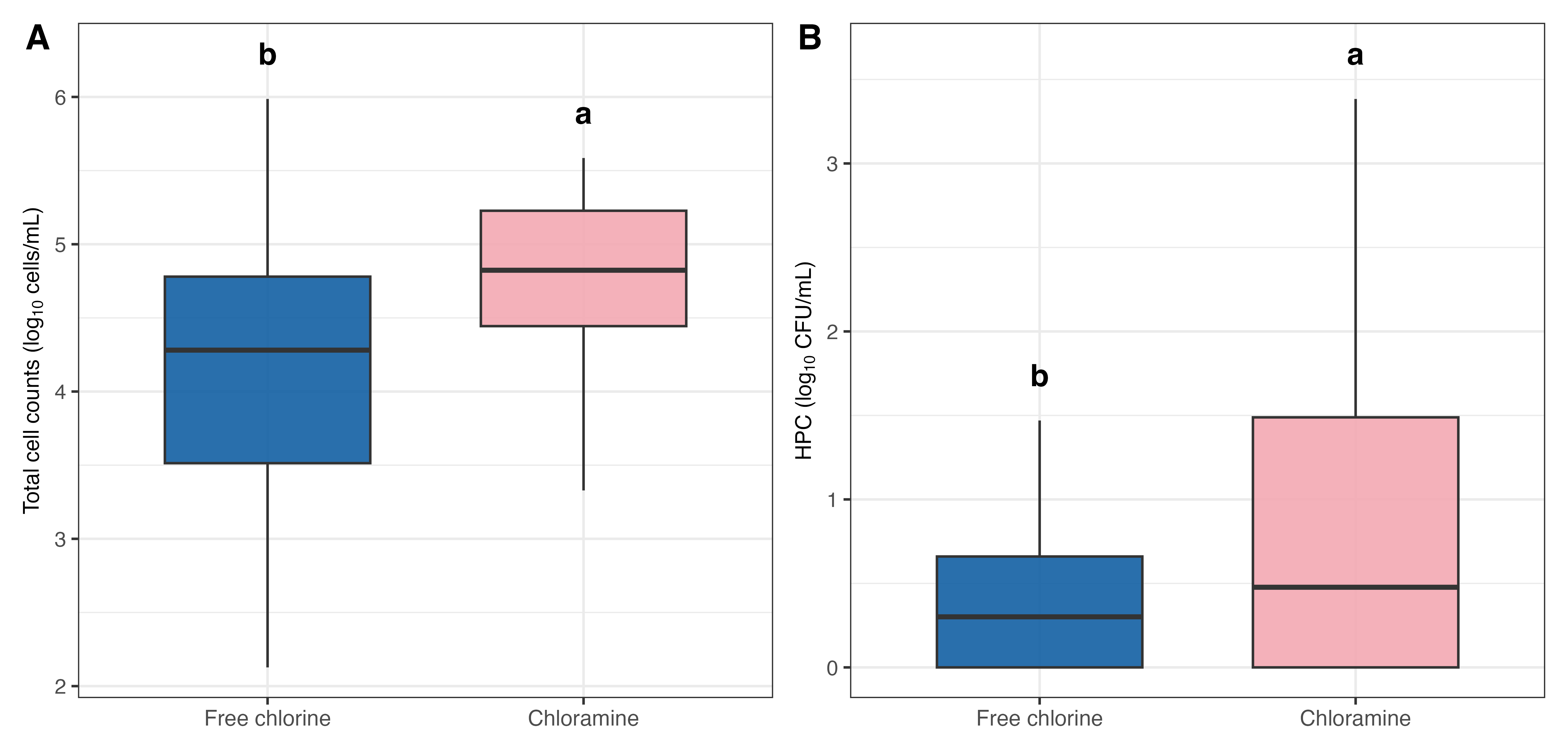

**Figure S6. Total cell counts and heterotrophic plate counts (HPC) between free chlorine and chloramine finished drinking water samples. (A) Total cell counts and (B) HPC. Different letters above boxplots denote statistically significant differences between disinfectant types (P < 0.05).**

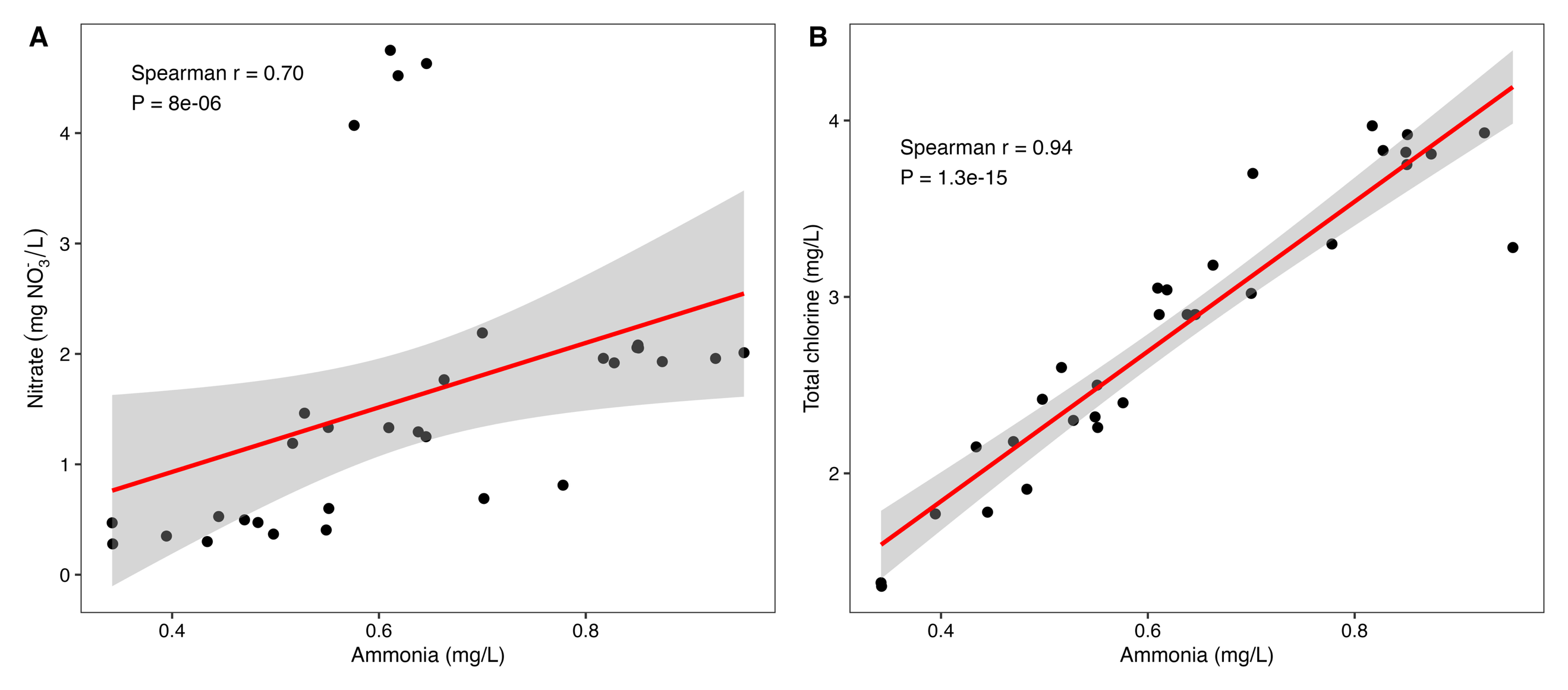

**Figure S7. Spearman correlation analysis between ammonia and (A) nitrate and (B) total chlorine. Red lines indicate fitted linear trends with 95% confidence intervals. Spearman’s correlation coefficient (r) and corresponding P value are shown in each panel.**

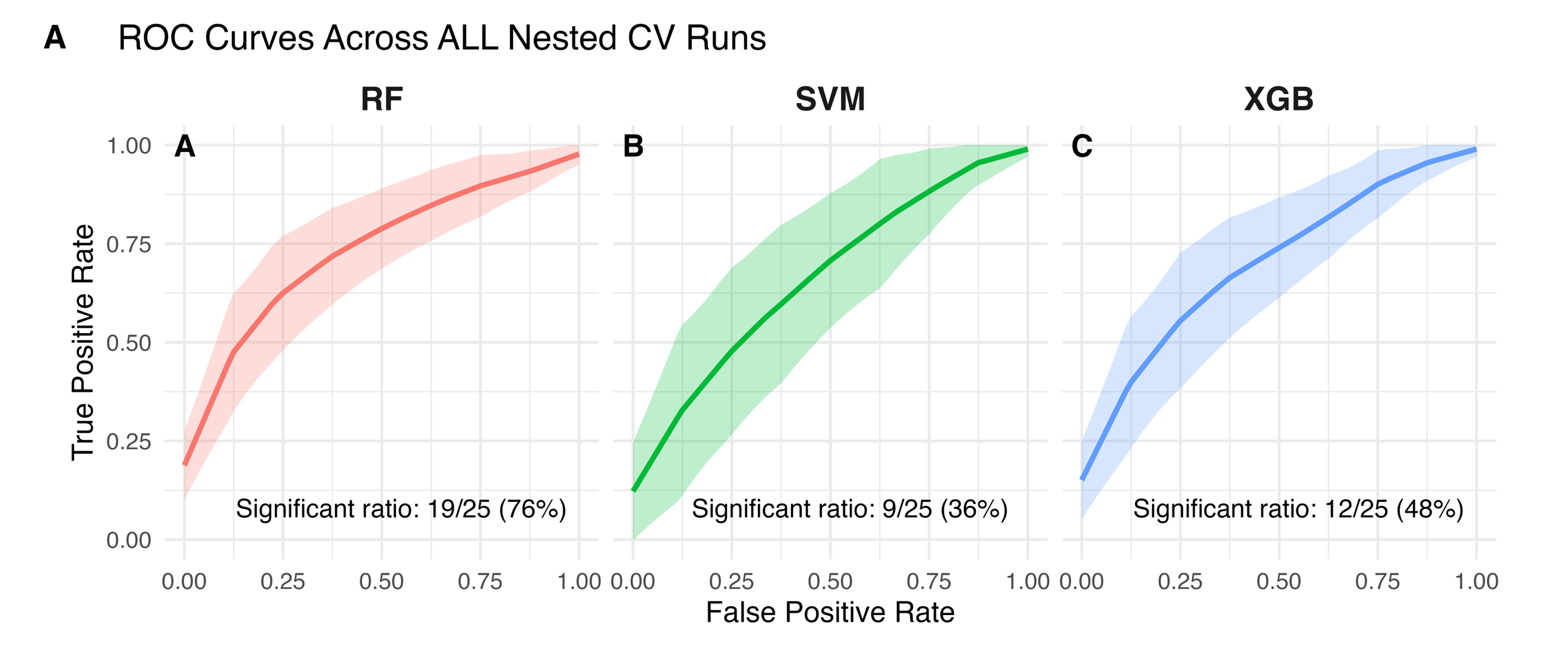

**Figure S8. Receiver operating characteristic curves (ROC) of Random Forest, Support Vector Machine, and Extreme Gradient Boosting models for predicting *Legionella* spp. occurrence across all nested cross-validation runs. Solid lines represent the mean true-positive rate across false-positive rates, and shaded bands indicate variability across runs. The significant ratio shown in each panel denotes the proportion of runs with statistically significant discrimination performance.**

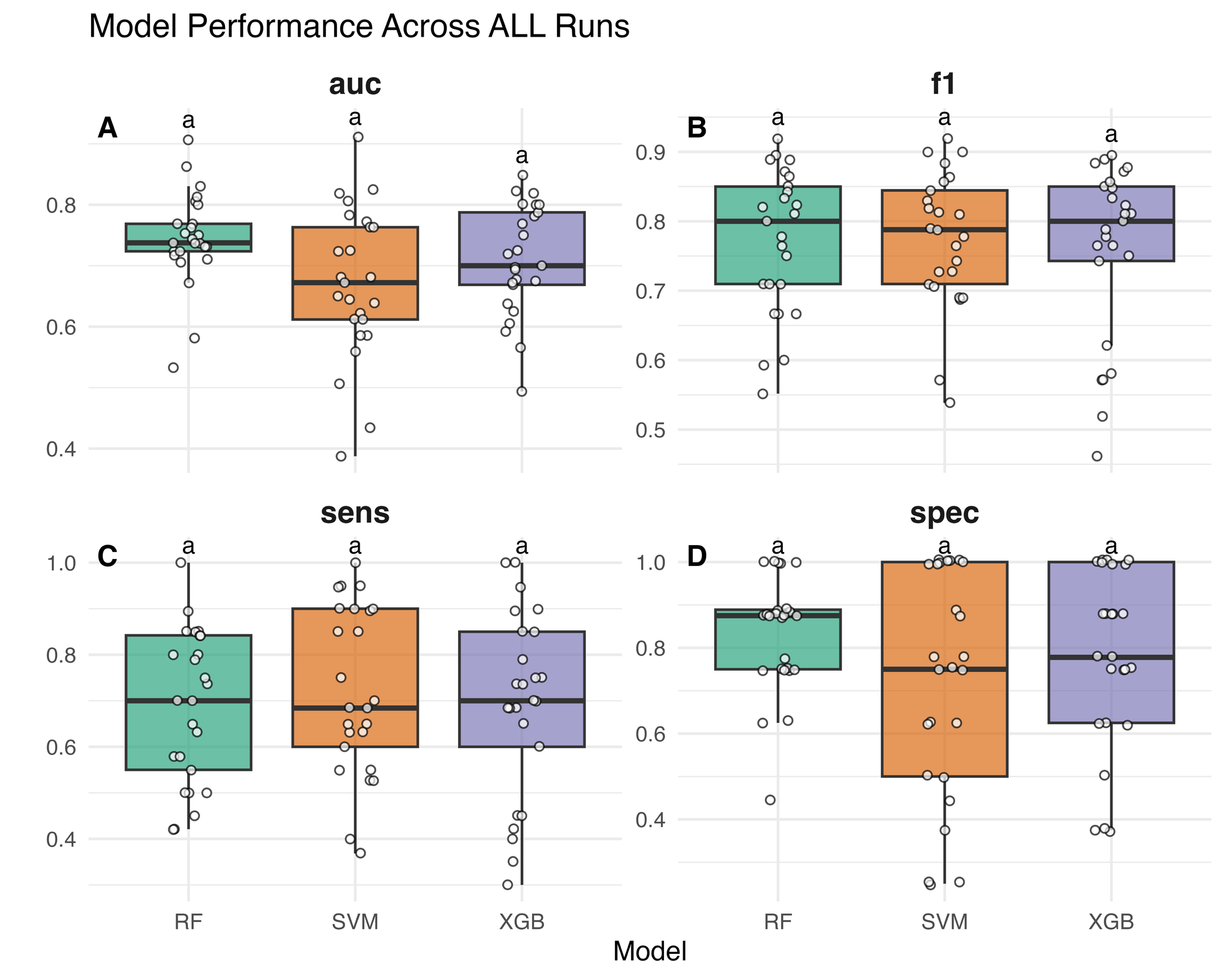

**Figure S9. Classification performance of Random Forest, Support Vector Machine, and Extreme Gradient Boosting models for predicting *Legionella* spp. occurrence across all nested cross-validation runs. Panels show distributions of area under the receiver operating characteristic curve (auc), F1 score, sensitivity, and specificity. Points represent individual cross-validation runs, and boxplots summarize the distribution of performance across runs. Different letters above boxplots denote statistically significant differences among models (P < 0.05).**

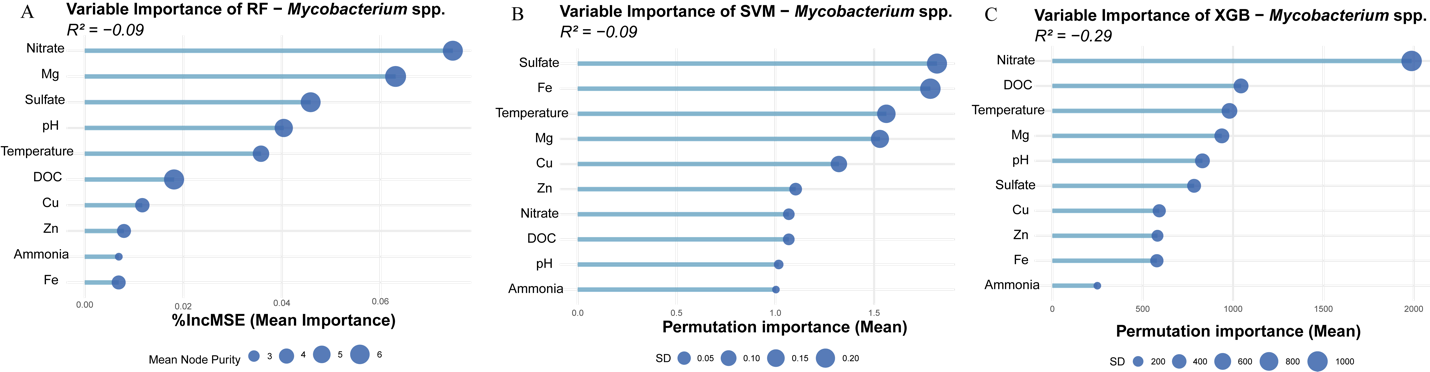

**Figure S10. Variable importance rankings for Random Forest, Support Vector Machine, and Extreme Gradient Boosting models predicting *Mycobacterium* spp. abundance. Panels show the relative importance of individual predictors in each model, ordered from highest to lowest contribution to prediction performance.**

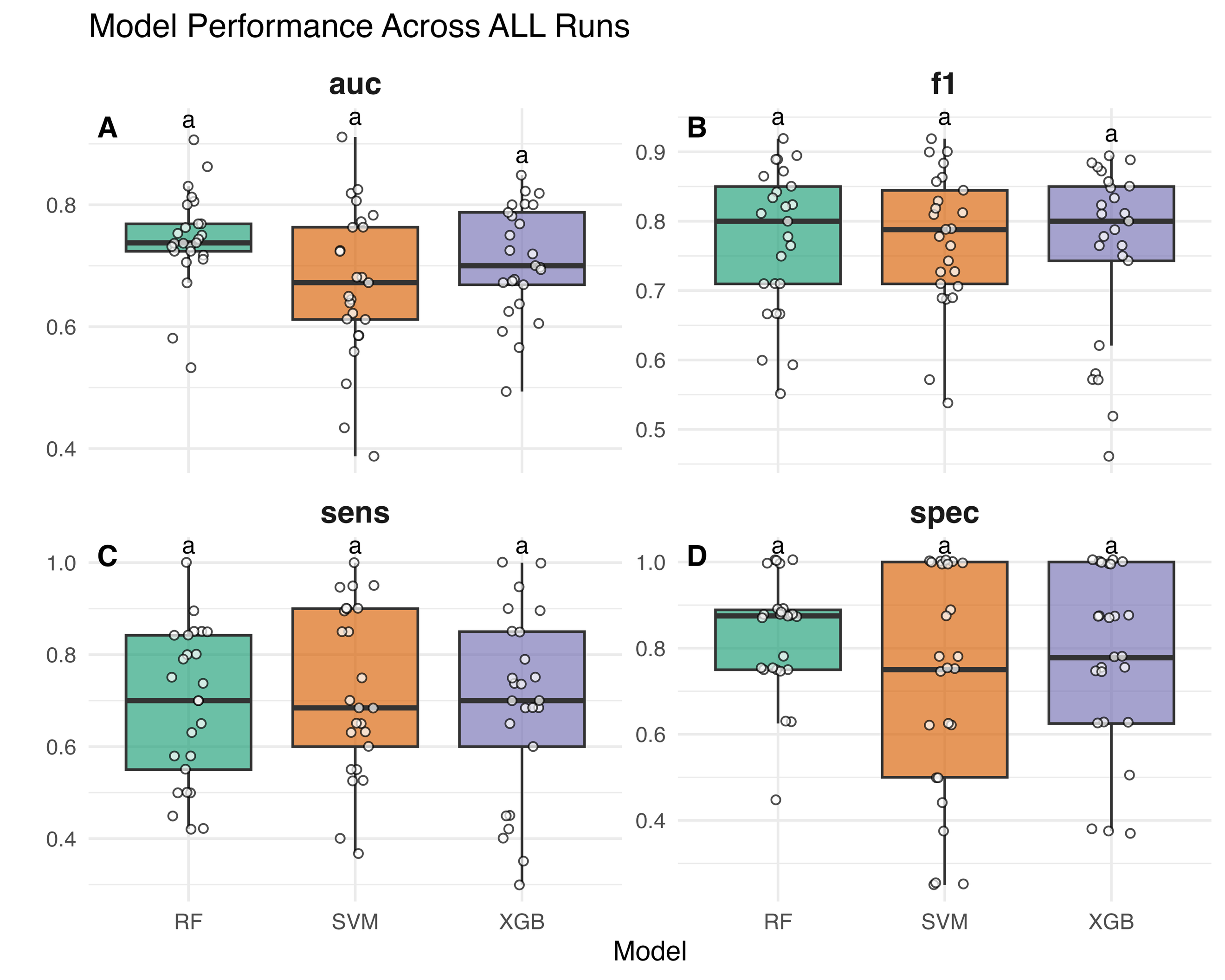

**Figure S11. Classification performance of Random Forest, Support Vector Machine, and Extreme Gradient Boosting models for predicting *Mycobacterium* spp. occurrence across all nested cross-validation runs. Panels show distributions of area under the receiver operating characteristic curve (auc), F1 score, sensitivity, and specificity. Points represent individual cross-validation runs, and boxplots summarize the distribution of performance across runs. Different letters above boxplots denote statistically significant differences among models (P < 0.05).**

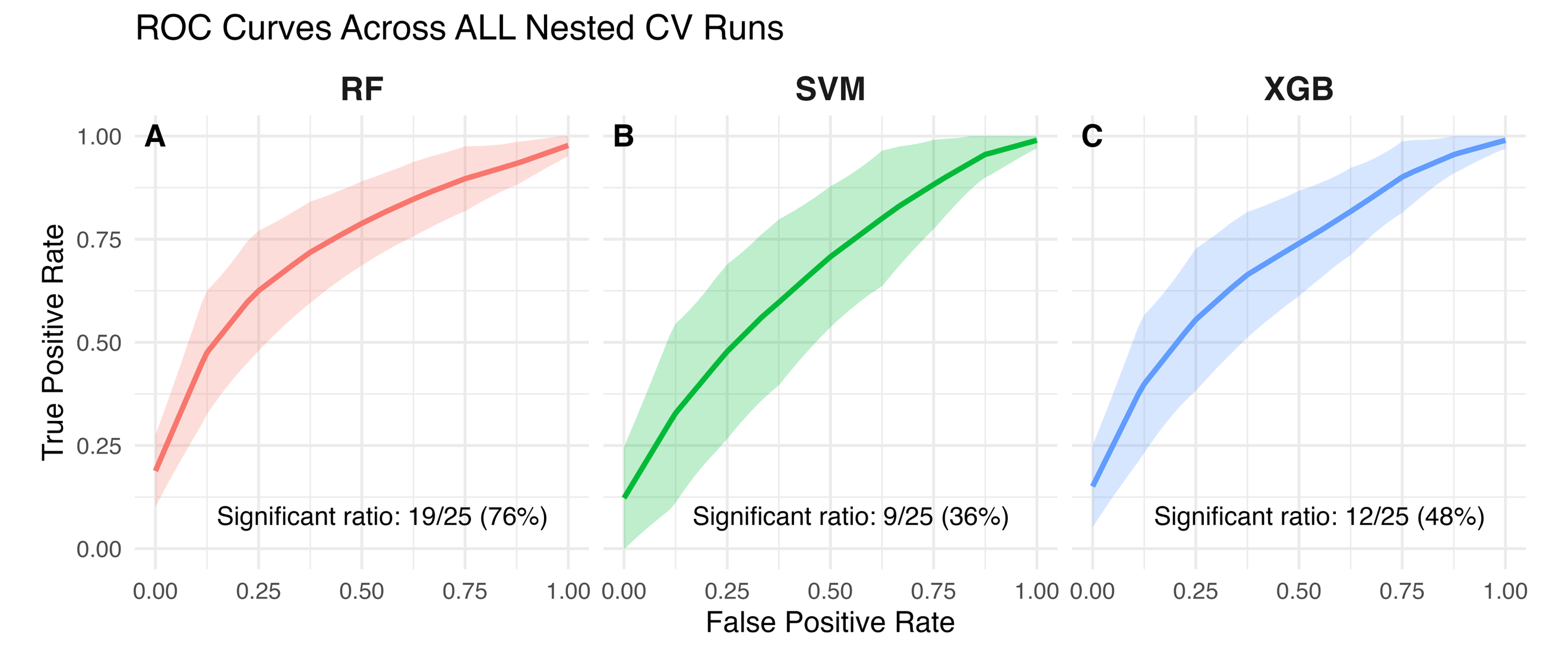

**Figure S12. Receiver operating characteristic curves (ROC) of Random Forest, Support Vector Machine, and Extreme Gradient Boosting models for predicting *Mycobacterium* spp. occurrence across all nested cross-validation runs. Solid lines represent the mean true-positive rate across false-positive rates, and shaded bands indicate variability across runs. The significant ratio shown in each panel denotes the proportion of runs with statistically significant discrimination performance.**
